# Propensity-score matching with GAN-generated observations from electronic health records: simulation study and application to the evaluation of prone positioning in COVID-19 patients under mechanical ventilation

**DOI:** 10.1101/2025.07.31.25332504

**Authors:** Bertrand Bouvarel, Benjamin Glemain, Fabrice Carrat, Nathanael Lapidus

**Affiliations:** Sorbonne Université, INSERM, Institut Pierre Louis d’Epidémiologie et de Santé Publique IPLESP, F75012 Paris, France; AP-HP.Sorbonne Université, Public Health Department, Saint-Antoine Hospital, F75012 Paris, France

**Keywords:** observational study, causal inference, propensity score matching, simulation, neural networks, generative adversarial networks, electronic health records

## Abstract

**Background:** Propensity score (PS) methods are widely used in observational studies to estimate causal effects, but they often exclude patients due to a lack of comparable counterparts, leading to reduced power and potential bias. Generative adversarial networks (GANs) have shown promise in creating synthetic data, but their application to causal inference remains underexplored. Synthetic data could be used as plausible counterfactuals, potentially mitigating the issues of the PS methods.

This study evaluates the integration of GAN-generated synthetic observations into propensity score matching (PSM) to improve the emulation of RCTs, using both simulated and real-world electronic health record (EHR) data.

**Methods:** A simulation study was conducted using with predefined confounding structures to compare traditional PSM against two hybrid approaches incorporating GAN-generated synthetic patients to partially or fully match the original sample of patients. Treatment effects were estimated via logistic regression, and performance was assessed by bias, standard error, alpha risk, power, and confidence interval coverage. The methods were applied to a real-world dataset of mechanically ventilated COVID-19 patients to evaluate the impact of early prone positioning on 28-day mortality.

**Results:** In simulations, GAN-generated patients permitted to match all patients in the original sample, whereas PSM dropped up to 60% of them. While synthetic augmentation improved sample size, unadjusted use of synthetic matches led to underestimated standard errors and inflated type I error. Down-weighting matched synthetic data improved error control but did not consistently outperform PSM in bias or power. In the real-world application (n=1399), treatment effect estimates for prone positioning were similar across all methods and did not reach statistical significance.

**Conclusion:** GAN-augmented propensity score matching can reduce sample loss. However, it’s current application in causal inference through PS matching remains limited. Synthetic data do not contribute independent information and must be cautiously integrated to avoid misleading precision. While promising, current GAN implementations require methodological refinements before routine use in causal inference.

## Background

Randomized controlled trials (RCTs) are regarded as the gold standard for evaluating interventions because randomization balances both measured and unmeasured confounders between treatment. [1] This balance permits causal attribution of outcome differences to the intervention. However, RCTs have well-known limitations. [2, 3] They are often expensive and time-consuming to conduct, especially for large populations or multiple sites. Ethical and practical constraints (e.g. the principle of equipoise) can preclude randomizing patients to certain exposures, particularly if early evidence or clinical judgment suggests one treatment might be superior. Additionally, RCT participants are usually managed under strict protocols in controlled settings, which may limit the generalizability of trial findings to broader real-world patient. [4]

In contrast, vast amounts of real-world data are now captured in electronic health records (EHRs). Over the past decade, EHR systems have enabled the aggregation of rich clinical data on millions of patients, including demographics, diagnoses, treatments, and outcomes. [5, 6] These EHR databases represent valuable resources for observational studies of treatment effects in diverse, routine-care populations. By analyzing EHR cohorts, researchers can address questions that might not be feasible in RCTs due to ethical or logistical reasons. [7, 8] However, because treatments in EHR data are not randomly assigned, observational analyses are vulnerable to confounding by indication and other biases. Patients receiving a given treatment may differ systematically from those who do not, and these differences (e.g., disease severity, comorbidities) can themselves affect outcomes, therefore simple comparisons of treated vs. untreated outcomes in such data can be very misleading, and such databases represent a valuable but yet scarcely used resource for the implementation of therapeutic evaluation.

Causal inference methods have been developed to explore causal hypotheses using observational data. [9] These methods are mainly based on counterfactual approaches via the construction of a propensity score (PS). [10] Two main PS-based strategies are frequently used: (1) inverse probability of treatment weighting (IPTW), [11] which reweights patients to create a pseudo-population with balanced covariates, and (2) propensity-score matching (PSM), [12, 13] which pairs treated and untreated patients with similar propensity scores and discards unmatched individuals. When properly applied, these techniques can mimic the randomized allocation of interventions that prevent confounding by indication in RCTs and reduce bias in estimated treatment effects from observational data.

The use of these methods with EHR data has already been proposed [14] using classical machine learning models. Despite the promising results of some studies, several limitations remain. Due to the nature of the data, in observational studies based on patients treated with knowledge of their symptoms/pathology, some patients may end up with a very high or low probability of receiving treatment (i.e., propensity scores very close to 0 or 1) because their clinical characteristics strongly predict treatment with very few, if any, counterfactual patients in other treatment groups. [15] Due to the violation of the positivity assumption, standard PSM must discard those patients due to lack of matches, with resultant power loss and selection bias, and IPTW must assign very large weights to a few rare patients, which inflates variance and leads to unstable estimates, especially when considering rarely-used treatments. [16]

One potential solution is to leverage modern machine learning to generate synthetic “counterfactual” patient data for the under-represented treatment group. In recent years, thanks to the improvement of algorithms and their support, neural networks have become increasingly popular for exploiting large databases [17, 18]. Among these networks, generative adversarial networks (GAN) are widely used thanks to their ability to generate realistic data. [19] A GAN consists of a *generator* network that produces artificial data (e.g. patient records) and a *discriminator* network that attempts to distinguish synthetic data from real data. [20] Through adversarial training, the generator learns to create increasingly lifelike outputs that the discriminator cannot easily detect as fake. GANs were initially popularized for image generation, but have since been applied to many domains including healthcare data. [21]

For example, researchers have used GANs to generate synthetic electronic health records and claims data that preserve important statistical properties of the originals. [22] Synthetic health data can facilitate data sharing and augment analyses while protecting patient privacy. [23] Specialized GAN architectures (e.g. medGAN, CTGAN) have been developed to handle tabular clinical data with mixed types (continuous, categorical). CTGAN in particular has demonstrated good performance in modeling complex multimodal distributions in patient records. [24] These advancements raise the question of whether GANs can be used to generate plausible counterfactual patients (e.g., a synthetic untreated version of a treated patient) to improve causal inference.

In this study, we propose to integrate GAN-generated synthetic observations into a propensity score matching framework and to study the impact on causal inference. We first conducted a simulation study to evaluate this approach under controlled conditions, examining whether matching with GAN data can recover known “true” treatment effects, in comparison with PSM. We then applied the method to a motivating example – the effect of early prone positioning on mortality in severe COVID-19 under mechanical ventilation – using EHR data from a large hospital network.

## Methods

### Simulation study

We designed a simulation study to compare traditional propensity-score matching with GAN-augmented matching methods. The overall workflow was as follows: (1) simulate “true” patient datasets with a known causal structure; (2) train a GAN on each dataset to generate synthetic patients; (3) perform different matching strategies (with and without synthetic data) to estimate the treatment effect; and (4) repeat across many simulations to evaluate performance.

#### Simulated data generation

Each simulated dataset consisted of 2000 patients with 8 baseline variables: 6 binary covariates representing patient characteristics (X_1_–X_6_), 1 binary treatment indicator (T) and 1 binary outcome (Y) for each hypothesis. Details about the simulation parameters are given in **Supplementary Text 1**.

We constructed the simulation so that treatment assignment was confounded by the baseline covariates, mimicking an observational study. Specifically, in *balanced* scenarios we set *P*(*T* = 1) = 50%, while in *unbalanced* scenarios *P*(*T* = 1) = 20%. The probability of treatment was modeled as a logistic function of certain covariates to introduce confounding by indication. We considered four underlying hypotheses for the data-generating process, depicted as directed acyclic graphs in **Figure 1**. Null hypotheses H0a and H0b considered that the treatment had no effect on the outcome. For H0a, the outcome was present with a 30% probability, independently of all other variables, whereas for H0b, the occurrence of the outcome depended on baseline characteristics.

**Figure 1.**
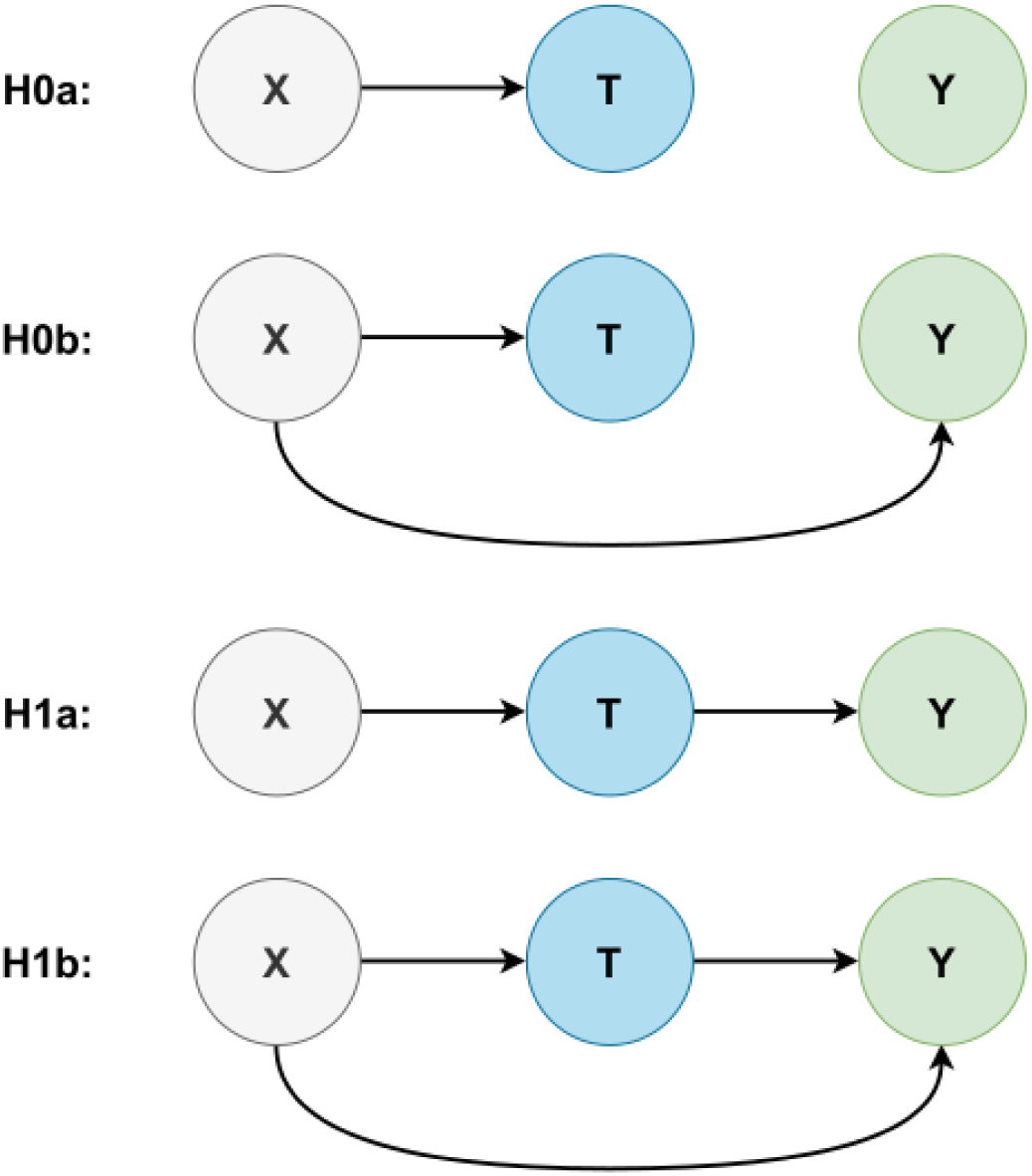
Directed acyclic graphs representing the 4 hypotheses of simulated causal associations. X: baseline characteristics of patients, T: treatment, Y: outcome.

Alternative hypotheses H1a and H1b considered that the treatment had an effect on the outcome (log odds ratio [OR] = 0.1), with H1b implying additional associations of the outcome with baseline characteristics. A sensitivity analysis was conducted with a higher effect size for H1a and H1b (log OR = 0.5). For each hypothesis, 1000 independent datasets were simulated.

*Note:* In this article, we distinguish “simulated” patients (those directly generated by our known simulation process) vs. “synthetic” ones (GAN-generated observations created from a simulated dataset).

#### GAN model for synthetic patient generation

After creating each simulated dataset, we used a GAN to generate additional synthetic patient records. Among considered architectures available for generating discrete or tabular data (mixture of discrete and continuous data), [25] we selected the CTGAN, specifically designed for tabular data and able to handle mixed continuous and categorical variables effectively. [24] [26] In our implementation, we simplified the data to binary features only, to ease the learning problem. We used the open-source Synthetic Data Vault (SDV) to train the CTGAN on each simulated dataset. [27] For each simulated dataset of 2,000 patients, we trained CTGAN until convergence (using default hyperparameters from SDV) and then generated a large pool of 50,000 synthetic patient records. This sizable pool was intended to include a wide variety of plausible patients drawn from the same underlying distribution as the original data. In particular, we expected that for most real patients in the dataset, one or more “counterfactual” synthetic patients could be found (i.e., a synthetic patient with very similar covariates but the opposite treatment value). We evaluated the fidelity of the GAN generation by comparing marginal and pairwise distributions between the simulated and synthetic samples.

#### Treatment effect estimation

The treatment effect on the outcome was estimated with univariable logistic regression. Propensity scores were constructed via multiple logistic regression, using baseline characteristics as predictors. The R MatchIt package [28] was used to match observations in the opposite treatment group and similar propensity scores, without replacement and using a caliper of 0.25. Three frameworks, depicted in **Figure 2**, were used to define the analyzed datasets:

– Propensity-score matching (PSM): construction of a PS in the initial simulated dataset and inclusion of pairs of treated/untreated patients with similar PS without relying on synthetic patients (any patient who did not have a match in the other treatment group was excluded);
– Partial hybrid matching (PHM): inclusion of the same pairs of simulated patients as for the PSM method, and addition of “hybrid pairs,” i.e. PS-matching of previously unmatched simulated patients with synthetic patients (PS in synthetic patients is calculated by applying to their baseline covariates the PS model derived from the simulated dataset);
– Full hybrid matching (FHM): PS-matching of all patients in the simulated dataset with synthetic patients, resulting in hybrid pairs exclusively.

**Figure 2.**
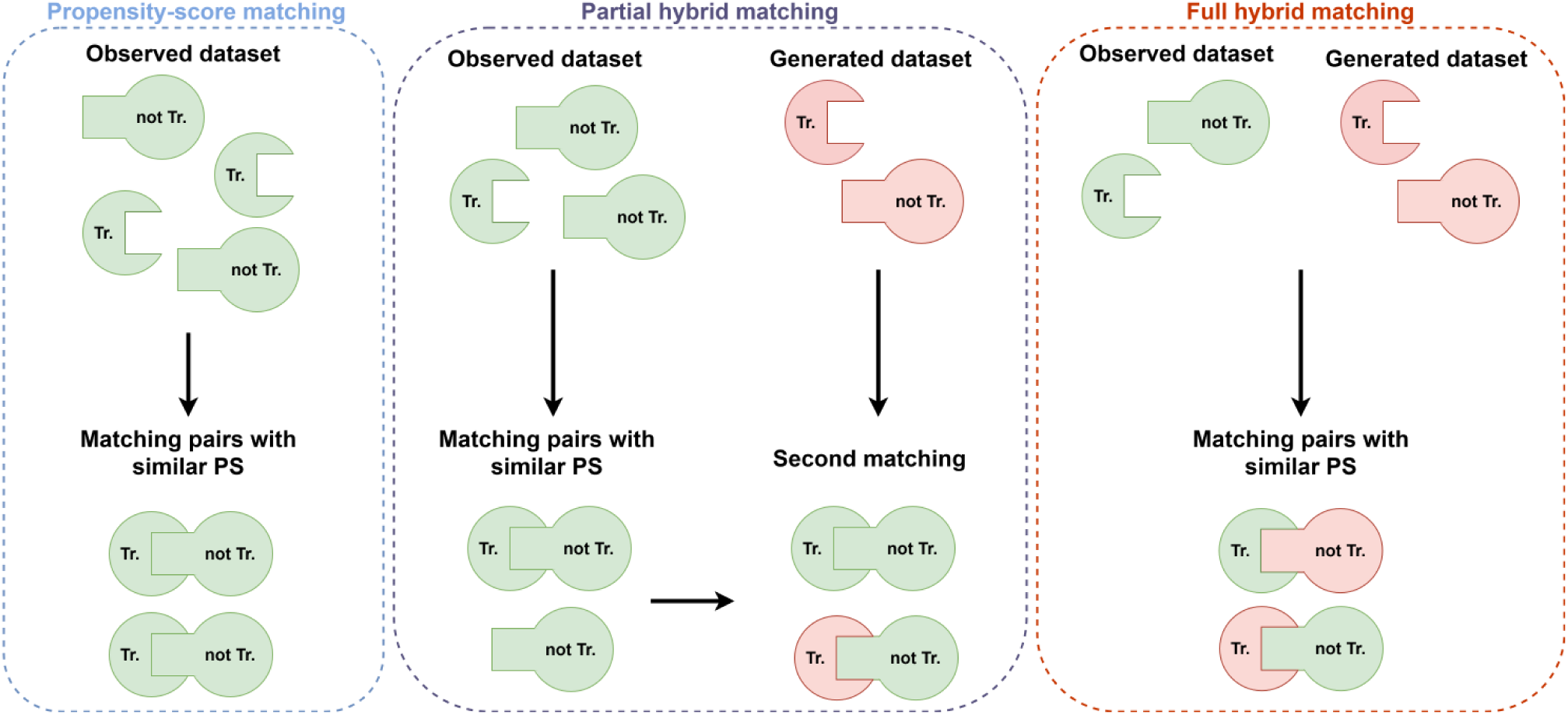
Diagram of the three frameworks of matching of the observation with similar propensity score. PS: propensity score, Tr.: treated, not Tr.: not treated.

Synthetic observations were generated using data from the simulated dataset and cannot be considered as independent from simulated observations. If treated as ordinary observations, the inclusion of such generated data would therefore increase the sample size without the additional information expected from independently sampled observations, and consequently underestimate the standard errors of log odds ratio (OR) in the FHM and PHM frameworks. To address this issue, we weighted the observations so that (i) the sum of weights in the analyzed dataset equaled the number of simulated observations, and (ii) both observations in a given pair had the same weight to avoid confounding by indication. Weights were therefore set to 0.5 for synthetic observations and their matched simulated observations, and 1 for all other observations. In a sensitivity analysis, weights were set to 0.25 for all synthetic observations and their matched simulated observations, with a sum of weights consequently lower than the number of simulated observations, whatever the number of synthetic observations.

#### Performance Metrics

The performances of these methods were assessed by evaluating their ability to correctly estimate the simulated treatment effect over 1000 simulated datasets. Judgment criteria were the bias (average difference between simulated and estimated log OR), the average standard error, the empirical standard error (standard deviation of the log OR point estimates), the type 1 error (alpha risk) and the statistical power (proportion of simulations with 95% confidence intervals (CIs) excluding 0 for null and alternative hypotheses, respectively), and the 95% CI coverage (proportion of 95% CIs containing the simulated effect) for alternative hypotheses. Additionally, we tracked the matched sample size (number of matched pairs) for each method. PSM drops patients to achieve balance, whereas PHM/FHM aim to retain more patients by using synthetic matches. We estimated how many more simulated patients could be kept by hybrid approaches.

### Motivating example: evaluation of prone positioning in COVID-19 mechanically-ventilated patients

Prone positioning – placing a patient in the face-down position – is a therapeutic intervention used in mechanically ventilated patients with acute respiratory distress syndrome (ARDS) in intensive care units (ICUs) to improve oxygenation. Its effectiveness in reducing mortality among patients with severe ARDS was demonstrated in a landmark randomized controlled trial in 2013. [29] Since then, further randomized investigations into its effectiveness in patient subgroups have been limited for ethical reasons, as its overall benefit in the ARDS population had already been established.

In this study, we applied the aforementioned methods to an observational EHR dataset to assess the effect of prone positioning on 28-day mortality in patients with severe ARDS caused by COVID-19. Specifically, we investigated whether GAN-augmented matching could lead to different conclusions or greater precision compared to standard propensity score matching. This analysis is compared to the target RCT it tried to emulate in **Supplementary Table 1**.

#### Study participants

Assistance Publique–Hôpitaux de Paris (AP-HP) is the university hospital trust operating in Grand Paris, receiving more than 10 million annual patient visits in 39 hospitals. A healthcare data warehouse (“Entrepôt de Données de Santé” AP-HP https://eds.aphp.fr) was set up in 2015 to conduct non-interventional research based on data collected during patients’ stays. [30] This database gathers clinical data, biology and microbiology tests, imaging data, prescriptions, textual data, as well as diagnoses coded with the International Classification of Diseases Version 10 (ICD-10) and medical procedures coded with the Classification commune des actes médicaux (CCAM). Data were extracted from this warehouse to retrieve all the patients who had performed a PCR test for SARS-CoV-2 with a validated result in any AP-HP hospital, which represents a dataset of more than one million patients.

Eligible patients were COVID-19 positives, as described in **Supplementary Table 1**, with respiratory distress requiring mechanical ventilation within 48 hours after ICU admission. Only patients admitted between March 1, 2020 and December 31, 2020 were selected (corresponding broadly to patients from the first and second waves, prior to vaccine availability). In this analysis, patients are defined as “treated” if prone positioning was initiated within 48 hours after ICU admission, whereas “control” patients were those without early prone positioning. Patients who died in the first 48 hours were excluded from the analysis, as we assumed that their critical state at admission would be an exclusion criterion from an RCT addressing the same question. The follow-up period started 48 hours after admission and ended after 28 days or death, whichever occurred first. A linkage of the data warehouse with the national database of deceased persons, provided by the National Institute of Statistics and Economic Studies (Insee), permitted assessment of vital status even after a patient was discharged from ICU. The judgment criterion for this analysis was 28-day mortality.

#### Variable selection and imputation

A broad set of variables available in the EHR data were selected to enter the analysis. These included risk factors commonly associated with COVID-19 morbidity and mortality, [31, 32] likely to be used for PS modeling, as well as miscellaneous variables describing patients’ conditions and characteristics, necessary for GAN generation. Those included biological measurements, medical procedures during hospitalization, or comorbidities, for a total of 50 variables. A detailed list of selected variables is provided in **Supplementary Table 2**, with their distribution in the treated and control groups. All variables were defined at baseline (within 48 hours following ICU admission).

Multiple imputation by chained equations was used to handle variables subject to missing values, using predictive mean matching for continuous variables and logistic regression for binary variables. [33, 34] Ten imputed datasets were created and independently used for synthetic patients generation and treatment effect estimation, using the previously described PSM, PHM and FHM frameworks. The distribution of selected variables after multiple imputation is reported in **Supplementary Table 3.** Continuous variables were binarized after imputation prior to data generation, to simplify the learning process of the CTGAN algorithm and avoid possible generated outliers, with threshold values chosen according to their deemed association with survival (**Supplementary Table 4**). The same estimation methods as those described in the simulation study were used to estimate the treatment effect in each imputed dataset. Treatment effect estimates (log OR) were finally pooled using Rubin’s rule.

Standardized differences were calculated to assess the balanced distribution of patient characteristics between intervention groups, with a threshold of 0.1 designated to indicate clinically meaningful imbalance. Positivity assumptions were assessed graphically by plotting the distribution of the propensity score in both intervention groups.

## Results

### Simulation study

#### Matched sample size

On average among 1000 simulations, the PSM results relied on approximately 711 pairs out of the 2000 simulated patients, i.e. a 28.9% loss of unmatched patients. The PHM and FHM frameworks, with balanced treatment allocation and 0.5 weights for patients in hybrid pairs, allowed to additionally match 577 simulated patients with synthetic ones, lessening the patients loss to 0.05% of the initial sample. This means a “counterfactual” generated patient could be found for almost all simulated patients, thus preserving the initial sample under all H0 and H1 hypotheses. With unbalanced treatment allocation, the patients loss for PSM was 60.2%, while PHM and FHM allowed to match most simulated patients with synthetic patients (0.015% and 0.01% loss, respectively).

#### Simulations under the null hypothesis

Results are reported in **Table 1**. Under the H0a and H0b hypotheses (absence and presence of confusion, respectively, with no treatment effect), the biases for usual PSM methods with balanced treatment allocation were –0.004 and 0.034, and the estimated alpha risks were 0.037 and 0.063, respectively. The biases for PHM were –0.002 and 0.035 under H0a and H0b, with alpha risks of 0.083 and 0.107, respectively. The biases for FHM were –0.003 and 0.039 for H0a and H0b, with alpha risks of 0.221 and 0.227, respectively. Compared with empirical standard errors, average standard errors were similar for PSM (0.116 vs. 0.113 and 0.133 vs. 0.140 under H0a and H0b, respectively), slightly lower for PHM (0.097 vs. 0.112 and 0.111 vs. 0.132, respectively) and lower for FHM (0.097 vs. 0.150 and 0.109 vs. 0.169, respectively). With unbalanced treatment allocation, the biases for PHM and FHM were higher than with balanced treatment allocation for H0a (0.042 and 0.036, respectively) and H0b (0.125 and 0.115, respectively), as well as the alpha risks. Compared with empirical standard errors, average standard errors were lower for PHM and FHM. Results regarding bias, alpha risk and comparison of average and empirical standard errors for PSM were similar to those with balanced treatment allocation.

**Table 1.**
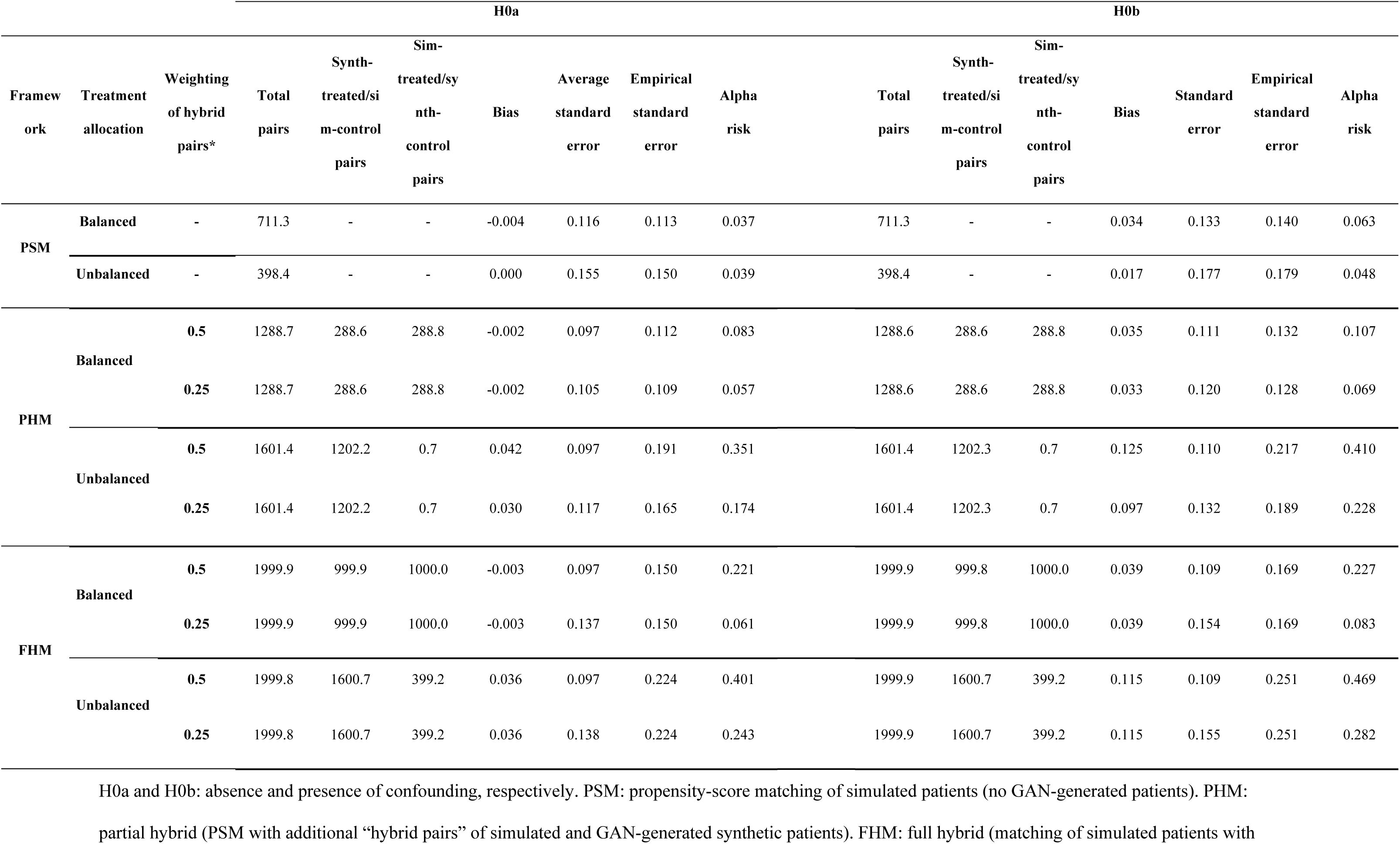
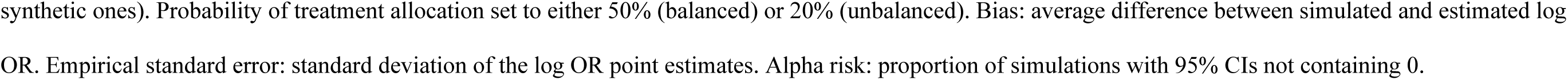
Performances of matching frameworks over 1000 simulations under the null hypotheses H0a and H0b.

Estimations with 0.25 weights for patients in hybrid pairs and balanced treatment allocation showed a better control of the alpha risk for both PHM (0.057 and 0.069 for H0a and H0b, respectively) and FHM (0.061 and 0.083, respectively). They also showed a lower difference between empirical standard errors and average standard errors for both PHM (0.105 vs. 0.109 and 0.120 vs. 0.128 under H0a and H0b, respectively) and FHM (0.137 vs. 0.150 and 0.154 vs. 0.169, respectively). Compared with the 0.5 weighting, the 0.25 weighting also showed a better control of the alpha risk with unbalanced treatment allocation for both PHM (0.174 and 0.228 for H0a and H0b, respectively) and FHM (0.243 and 0.282, respectively), as well as a lower difference between average and empirical standard errors for both frameworks.

H0a and H0b: absence and presence of confounding, respectively. PSM: propensity-score matching of simulated patients (no GAN-generated patients). PHM: partial hybrid (PSM with additional “hybrid pairs” of simulated and GAN-generated synthetic patients). FHM: full hybrid (matching of simulated patients with synthetic ones). Probability of treatment allocation set to either 50% (balanced) or 20% (unbalanced). Bias: average difference between simulated and estimated log OR. Empirical standard error: standard deviation of the log OR point estimates. Alpha risk: proportion of simulations with 95% CIs not containing 0.

#### Simulations under the alternative hypothesis

Results are reported in **Table 2**. Under the H1a and H1b hypotheses (absence and presence of confusion, respectively, with a treatment effect of log OR = 0.1), the biases for usual PSM methods with balanced treatment allocation were –0.006 and 0.028, and the estimated powers were 0.121 and 0.096, respectively. Using 0.5 weights for patients in hybrid pairs, the biases were similar for PHM (–0.005 and 0.028, respectively), as well as for FHM (–0.009 and 0.038, respectively). Compared with PSM, powers were higher for PHM (0.194 and 0.140, respectively) and FHM (0.302 and 0.225, respectively). PSM showed appropriate 95% CI coverage (95.3% and 94%, respectively), while it was close for PHM (91.1% and 90.8%, respectively) and lower for FHM (79.5% and 79.6%, respectively). With unbalanced treatment allocation, performances were similar for PSM under both H1a and H1b hypotheses, whereas bias and 95% CI coverage showed poorer performances for both PHM and FHM.

**Table 2.**
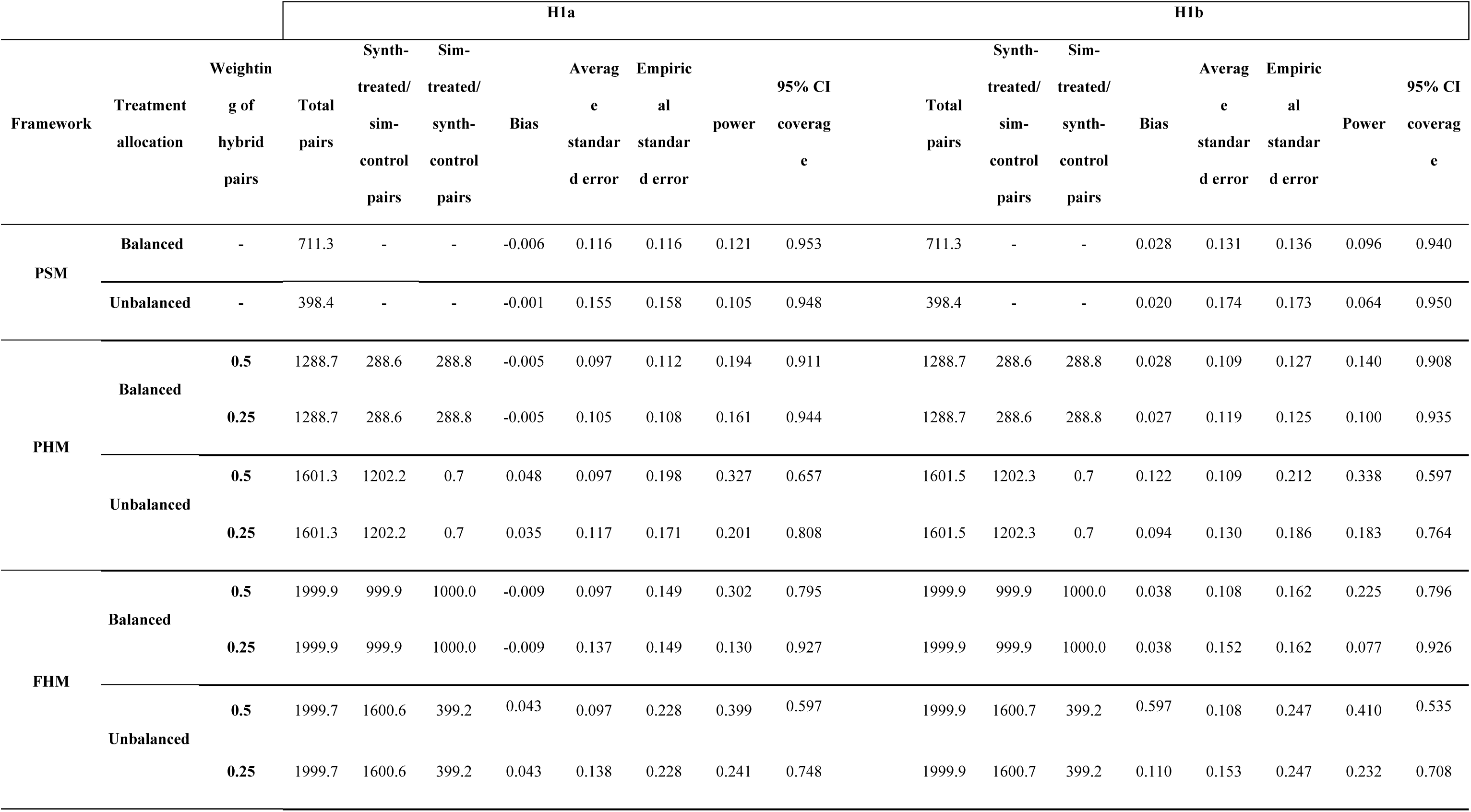
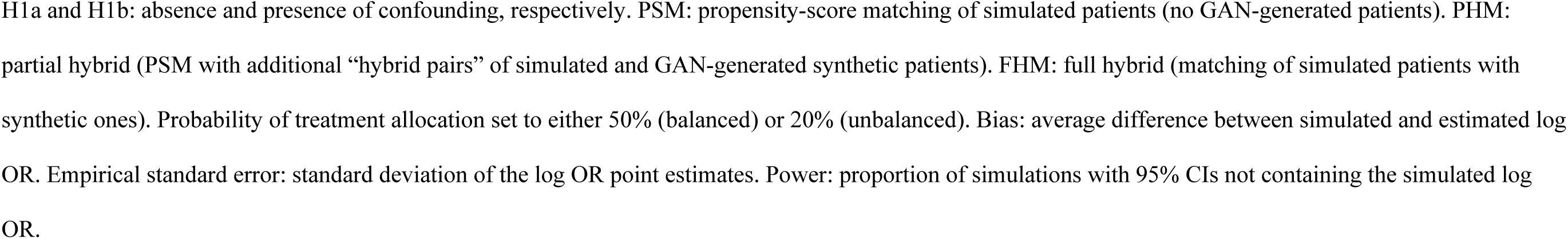
Performances of matching frameworks over 1000 simulations under the alternative hypotheses with simulated log OR = 0.1.

Estimations with 0.25 weights for patients in hybrid pairs showed similar or better performances than the 0.5 weighting regarding all performance measures for both PHM and FHM, with either balanced or unbalanced treatment allocation. With balanced treatment allocation, 95% CI coverages were close to the expected 95% value for both PHM (94.4% and 93.5% under H1a and H1b, respectively) and FHM (92.7% and 92.6%, respectively).

When simulating a log OR of 0.5, the comparison of frameworks under the different scenarios led to similar conclusions, with performances globally poorer than with log OR = 0.1 (**Supplementary Table 5**).

### Evaluation of prone positioning in COVID-19 mechanically-ventilated patients

#### Study participants

A total of 1459 unique COVID-19 patients met the inclusion criteria, among whom 60 had unknown discharge mode. In the 1399 remaining patients, 590 belonged to the prone positioning (treated) group and 809 to the control group. The number of deaths in these groups were 226 and 254, respectively (**Figure 3**). Baseline characteristics of study participants are reported in **Supplementary Table 2.** The median age was 63 years (Q1: 55; Q3: 70), 75% were male, and common comorbidities included history of cancer (6%), hypertension (34%), renal disease (33%), and diabetes (24%). By study design, all patients were on mechanical ventilation for acute respiratory failure.

**Figure 3.**
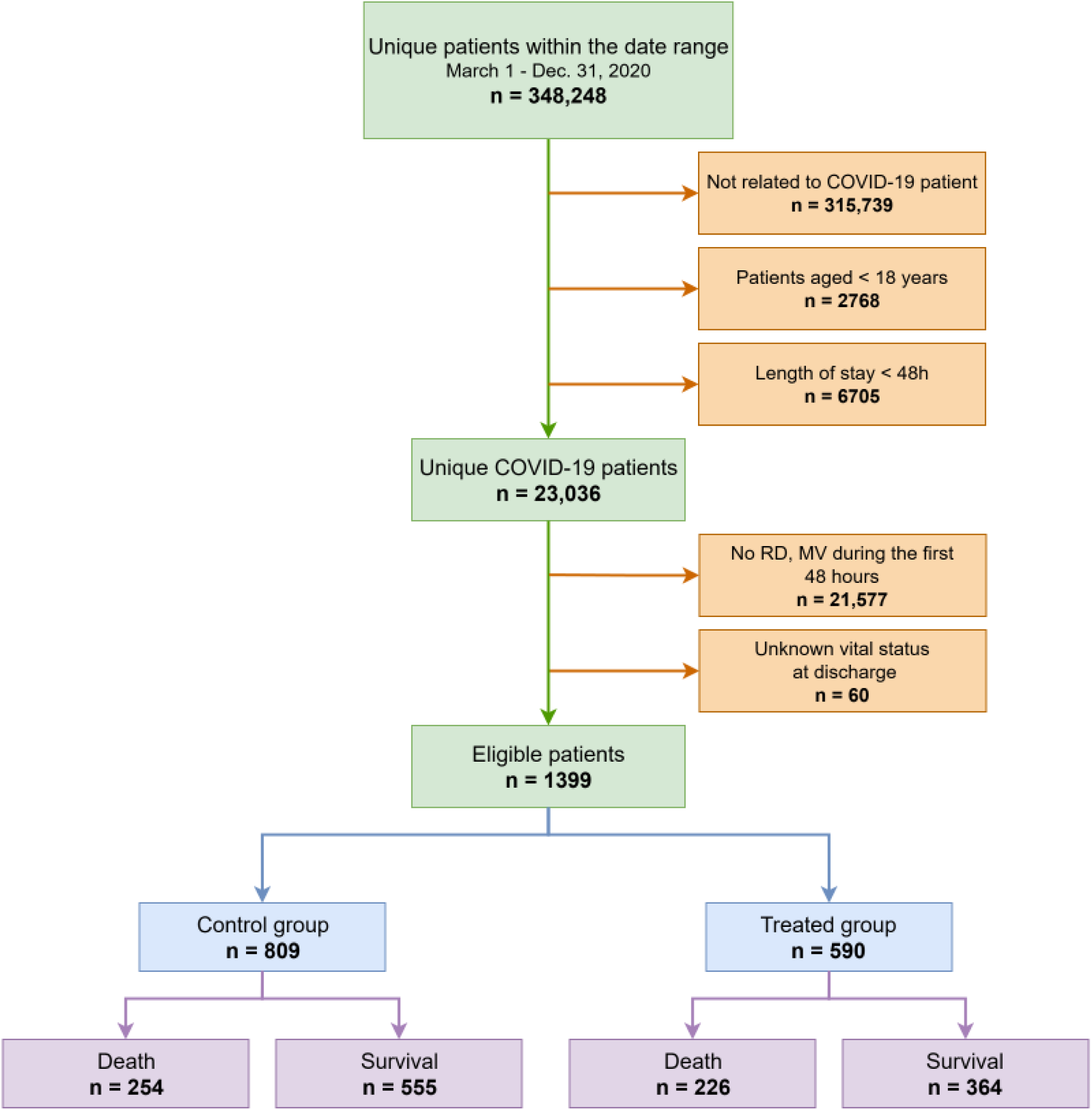
Flowchart for the inclusion and survival of patients from the AP-HP warehouse. RD: respiratory distress, MV: mechanical ventilation.

#### Data generation

Multiple imputation was performed over the 50 baseline covariates (averaged values over the 10 imputed datasets are reported in **Supplementary Table 3**). After continuous covariates were binarized, the 10 imputed datasets were independently submitted to the CTGAN algorithm to generate 50,000 synthetic patients.

#### Quality control

Intra-variable quality, i.e. whether the model was able to reproduce the marginal distribution of each variable in the initial cohort, was checked by graphically comparing the means and standard deviations between observed and generated data (**Supplementary** Figure 1). The distribution of generated values was very similar to observed data, except for renal replacement therapy (RRT), absent in all generated patients.

**Supplementary** Figure 2 shows the cumulative sum of variables in observed and generated patients, thus providing a more detailed overview of marginal distributions.

Joint distributions were checked graphically by comparing the correlograms between observed and generated data (**Supplementary** Figure 3). As for marginal distributions, observed and generated data showed very similar patterns of associations, except for RRT.

#### Matching

The same matching frameworks as those evaluated in the simulation study were used. PSM with a 0.25 caliper allowed to match an average of 471.5 patients (80.0% of patients in the treated group and 58.3% of the control group). Both PHM and FHM were able to match all the 1399 observed patients, using generated patients (**Table 3**). **Figure 4** compares the distribution of propensity scores between the treated and control groups before matching and using PSM, PHM and FHM. The use of hybrid pairs helped attain balance in measured baseline covariates between treated and control subjects, with only 27 standard mean differences out of 63 below the 0.1 threshold without matching, vs. 14, 11 and 7 after PSM, PHM and FHM, respectively (**Supplementary** Figure 4).

**Figure 4.**
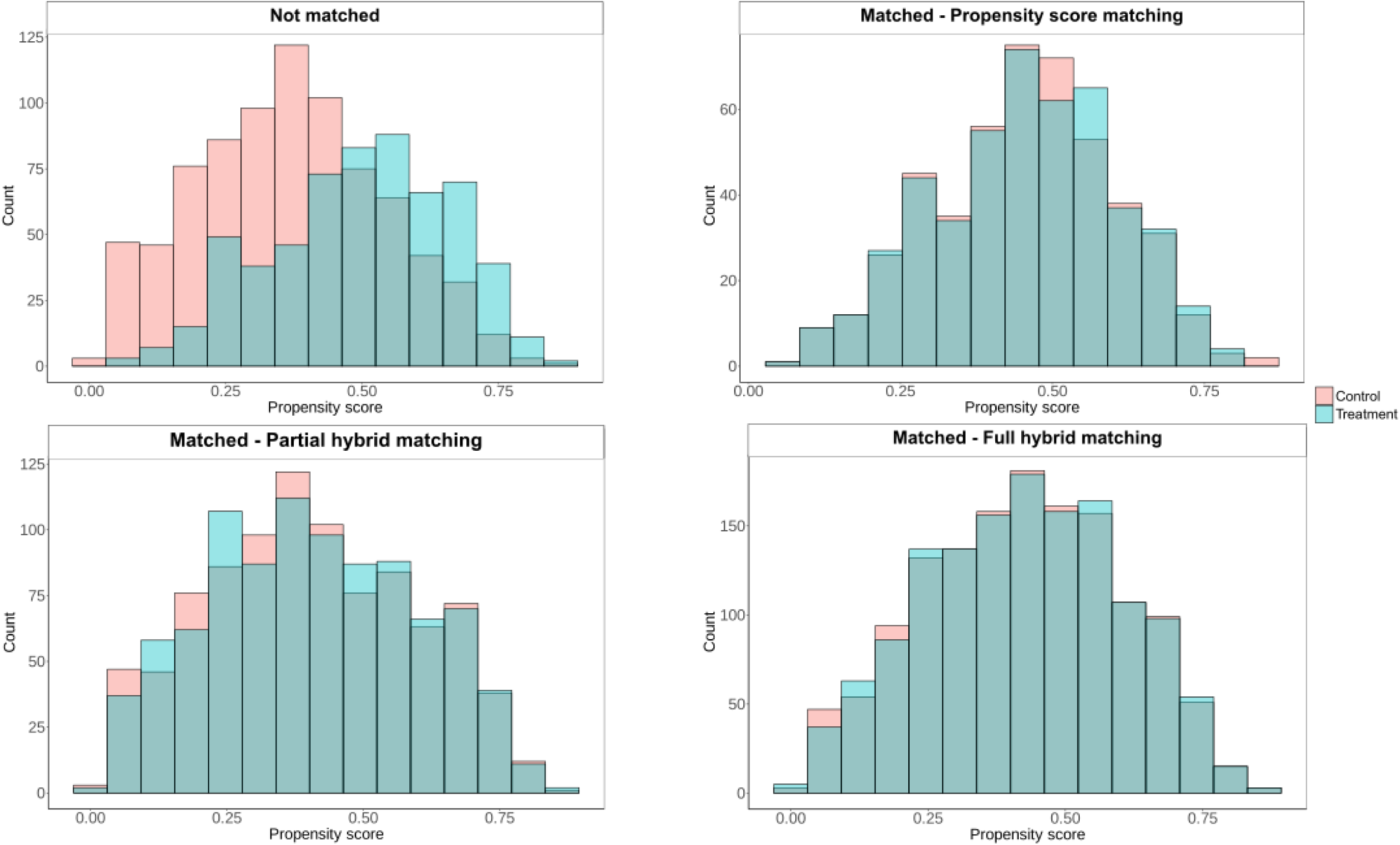
Histogram of propensity scores for the two treatment groups before and after matching.

**Table 3.**
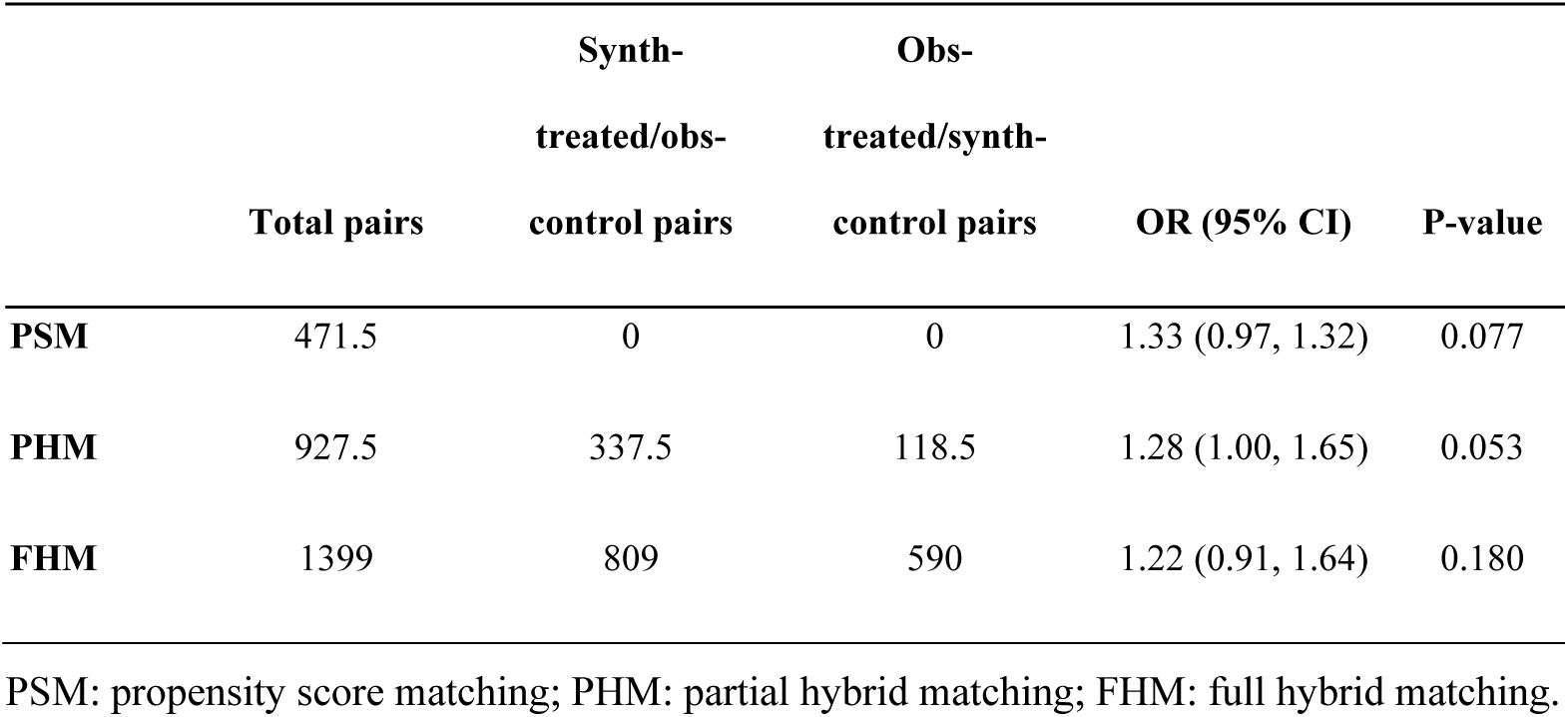
Estimation of the association of prone positioning with 28-day mortality of COVID-19 patients in respiratory distress.

#### Effect estimation

The OR point estimates for the effect of prone positioning on 28-day mortality were similar for all matching frameworks: 1.33 (0.97, 1.32), 1.28 (1.00, 1.65) and 1.22 (0.91, 1.64) for PSM, PHM and FHM, respectively (**Table 3**). No association was significant at the 0.05 threshold.

## Discussion

### Main findings

In this work, we explored a novel approach to augment data in causal inference analyses with GAN-generated synthetic observations. The motivation was to address a common shortcoming of PS matching methods: the exclusion of patients without matches in the other intervention group. By generating synthetic patients for the under-represented strata, we hoped to include as many patients as possible from the original sample in the analysis, thus potentially increasing statistical power and generalizability.

Our simulation results indicate that, while GANs can indeed create very realistic synthetic patients and prevent sample loss, the current implementation did not reliably improve causal inference performance. In fact, naively using all GAN-generated matches led to misleadingly small p-values standard deviations and confidence intervals, i.e. an anti-conservative analysis. This was evidenced by the inflated type I error under null scenarios and under-coverage of confidence intervals under alternatives. The fundamental issue is that synthetic data do not add independent information – they are derived from the original data and essentially replicate patterns from it. If treated as entirely new observations, they cause the analysis to overcount evidence, resulting in overstated precision. Down-weighting pairs with synthetic observations limited the type I error, yet no scenario permitted to obtain lower type I and type II error rates than PSM simultaneously.

The real-world application echoed these findings. In the COVID-19 prone positioning study, PSM excluded a large number of patients (in this case, many controls who had much lower risk and thus were unmatched). PHM was able to include nearly all patients by pairing them with GAN-synthetic counterparts, roughly doubling the matched sample size. However, the treatment effect estimates remained essentially unchanged and there was still no statistically significant effect of prone positioning on mortality with either approach. FHM, which used exclusively synthetic matching, did not improve precision at all in this case. We suspect this is because the GAN introduced noise and slight distortions in the data that offset the gains from augmented sample size. Indeed, our GAN had excellent performance in reproducing marginal distributions, but subtle discrepancies in joint distributions or extreme values could have diluted the treatment contrast. Importantly, even if PHM had yielded a p-value <0.05, one would need to be cautious in interpretation, as the nominal significance might be driven by underestimated uncertainty.

Other differences between the three frameworks might also be explained by the estimators: in causal inference, the average treatment effect (ATE) estimates the average impact of a treatment across the entire population, regardless of who actually received it. In contrast, the average treatment effect on the treated (ATT) focuses only on those who were treated, measuring the treatment’s effect specifically for them. By discarding control patients useless to match treated ones, PSM usually follows an ATT approach (when most patients in the sample are not treated). By considering all patients in the original sample, hybrid methods are closer to an ATE approach. We expected this difference would reveal the interest of hybrid methods in the unbalanced scenario, as the gain in sample size was the highest, yet results were quite similar to the balanced scenario.

### Challenges of augmented data in causal inference

Our study highlights several challenges and considerations for future efforts. First, regarding variance estimation: The most prominent issue is how to correctly estimate uncertainty when synthetic data are used. Standard formulas assume independent samples; here that assumption is violated. We applied a simple weighting heuristic as a proxy for more formal variance correction; alternative approaches could involve analytical derivation of the effective sample size of GAN-augmented data or using bootstrapping techniques that resample the original data and regenerate synthetic data in each bootstrap, which would be computationally heavy. Without adjustment, one risks reporting overly tight confidence intervals and exaggerated significance. Any practical application of GANs in inference must address this, possibly by scaling standard errors upward or using sandwich estimators that account for the two-stage generation process.

Second, regarding the bias vs. variance trade-off: Interestingly, we did not observe notable systematic bias in point estimates due to GAN usage. However, one can imagine scenarios where the GAN could introduce bias: for instance, if the GAN fails to capture an interaction between treatment and a covariate that affects outcome, the synthetic data might distort the effect estimate. Third, regarding GAN limitations: The performance of this strategy ultimately depends on the quality of the GAN-generated data. If the GAN perfectly reproduced the joint distribution of covariates and the treatment assignment mechanism, then synthetic controls would be as reliable as real ones (assuming no unmeasured confounding). However, current GANs have limitations. Although their use for image generation is showing very satisfactory results, [35, 36] our results highlight their limits for discrete and tabular data, which corroborates recent research on the architecture we used. [37] They may struggle with high-dimensional, sparse data or with capturing complex correlations. For example, generating a realistic combination of multiple comorbidities and lab trends that correspond to a severe COVID patient is a tall order. Our chosen model, CTGAN, is state-of-the-art for tabular data, but even it might not encode all clinical logic. GANs can also suffer from mode collapse (not representing the full diversity of the data) or can produce nonsensical outliers. In our synthetic data checks, we noticed a few minor inconsistencies (like a synthetic patient with an unusual combination of rare features that was not seen in reals, but these were rare). If GANs inadvertently violate clinical constraints or logical relationships, those could confound the matching. Recent studies concur that generating complex, multimodal health records with full fidelity is an open challenge. [22] For instance, Ghosheh et al. point out that current GAN approaches cannot yet generate richly detailed patient records across multiple data types (labs, images, notes) in a coherent way. [38] In our context, we only dealt with tabular ICU data and still faced difficulties. Thus, applying GANs in more complicated causal questions (e.g. longitudinal treatments, time-varying confounding) will be even more challenging.

Fourth, regarding generative model improvements: There is active research into better generative models for healthcare data. Our negative findings don’t mean GANs will never be useful – rather, it highlights that methodological advances are needed. One avenue is to incorporate domain knowledge or constraints into GAN training. Integrating clinical rules could prevent unrealistic combinations. [22] This could be achieved via constrained GAN objectives or post-processing filters. Additionally, non-GAN generative methods like Bayesian models or imputation models could be explored as alternatives for generating counterfactuals. Regression models [39], CART models that have recently shown more promising results than current GANs [37], or transformers fine-tuned for tabular data [40] may be considered. Other approaches might avoid some pitfalls of GANs but have their own modeling assumptions. [41]

Fifth, regarding the use of synthetic data as external control arms: The idea of using external or synthetic controls is gaining traction in regulatory settings (e.g. single-arm trials augmented by real-world data controls). [23] The experience here aligns with cautions raised in that context: quality and comparability of the external data (or synthetic data) are paramount. Regulators emphasize thorough validation of whether an external control truly “mirrors” a randomized control. In our case, we see that even though our GAN synthetic patients passed basic similarity checks, using them required statistical adjustment to avoid bias. This reinforces that synthetic data should be used with great care in causal analyses, and methods to quantify uncertainties are needed.

### Limitations

This study has several limitations. First, our simulation scenarios, while diverse, may not encompass all situations. We used relatively low-dimensional data (six covariates) and simple logistic models. In more complex simulations (e.g. nonlinear effects, high-dimensional covariate space, presence of interactions), the performance of the evaluated matching frameworks might differ. Second, we focused on one GAN model (CTGAN). It is possible that other generative models (e.g. variational autoencoders, normalizing flows, or newer transformer-based generators) could produce higher fidelity synthetic data and improve results. Third, in the prone positioning case study, there may be unmeasured confounders that no amount of GAN matching can fix. Our methods, like any PS method, assume no residual confounding, whereas the prone positioning example is complicated by indication bias not captured by the documented covariates. Thus, one must remember that synthetic data augmentation addresses data overlap and sample size issues, not fundamental confounding. Finally, our weighting strategy for synthetic data, while pragmatic, lacks a rigorous theoretical foundation and an optimal weighting or a different variance estimator might be derived in future work.

### Future research

We encourage further methodological research on how to integrate augmented data into causal analyses. For instance, developing causal inference algorithms that internally generate synthetic counterfactuals and adjust for their uncertainty could be fruitful. There is ongoing work on bridging GANs with causal modeling – some propose training GANs not just to match data distribution, but also to ensure that causal relationships (e.g. conditional independencies, propensity score distributions) are preserved. [42] Additionally, the use of large language models (LLMs) and transformers for data synthesis is emerging as an alternative to GANs. Early evidence suggests LLMs might generate mixed-type data with fewer mode collapse issues, which could potentially improve tabular data synthesis quality. [22] From an application perspective, more case studies in different clinical settings might be useful. Scenarios with major indication bias and limited sample size may uncover situations where the advantages of data augmentation might overcome its drawbacks.

## Conclusions

Our study provides a cautious take on the use of GAN-generated synthetic data for causal inference in medicine. GANs show great promise in creating realistic patient data and can help keep more patients in causal inference with propensity score matching. However, simply adding GAN-generated patients is not recommended without careful adjustments, as it can lead to erroneous confidence in results. In our simulations, standard PSM still provided more reliable control of type I error and coverage than unadjusted hybrid approaches. We advise that researchers proceed carefully when considering synthetic data augmentation: validate that synthetic records are indistinguishable from real in key covariates, ensure balance on propensity scores is achieved, and adjust the analysis to account for the nature of the generated data. As generative AI techniques evolve, we anticipate they will become more robust and perhaps eventually play a routine role in observational studies. However, the use of GANs in causal inference still requires skepticism and rigorous assessment, to which our work contributes by highlighting pitfalls and suggesting practical solutions. With continued research at the intersection of machine learning and causal inference, we remain optimistic that future innovations will unlock the full potential of synthetic data while maintaining the rigor of causal analysis.

## Declarations

### Ethics approval and consent to participate

This multicenter, non-interventional research is based on care data collected during patients’ hospital stays at AP-HP (reference: 200016 PREDIREA-COVID). It was approved by the Scientific and Ethics Committee of the AP-HP Health Data Warehouse (EDS), which is authorized by the French National Commission on Informatics and Liberty (CNIL) to review such non-interventional, data-based studies that do not require informed consent. The research does not involve processing indirectly identifiable data, linking with external data sources, or conducting long-term patient follow-up.

### Consent for publication

Not applicable.

### Availability of data and materials

The data used in the study are not publicly available due the confidentiality of data from patient records, even after de-identification. However, access to the AP-HP data warehouse’s raw data can be granted following the process described on its website www.eds.aphp.fr, contacting the Scientific and Ethics Commity at. A prior validation of the access by the local institutional review board is required. In the case of non-APHP researchers, the signature of a collaboration contract is moreover mandatory.

### Authors’ contributions

BB, BG, FC and NL designed the study. BB and NL conducted the analyses and drafted the manuscript. All authors revised it critically and approved the final version.

## Data Availability

The data used in the study are not publicly available due the confidentiality of data from patient records, even after de-identification. However, access to the AP-HP data warehouse's raw data can be granted following the process described on its website www.eds.aphp.fr, contacting the Ethical and Scientific Commity at. A prior validation of the access by the local institutional review board is required. In the case of non-APHP researchers, the signature of a collaboration contract is moreover mandatory.

## List of abbreviations

RCT: Randomized Controlled Trial
EHR: Electronic health record
PS: Propensity Score
IPTW: Inverse Probability of Treatment Weighting
PSM: Propensity Score Matching
GAN: Generative Adversarial Network
CTGAN: Conditional Tabular GAN
SDV: Synthetic Data Vault
PHM: Partial Hybrid Matching
FHM: Full Hybrid Matching
CI: Confidence Interval
OR: Odds Ratio
ARDS: Acute Respiratory Distress Syndrome
ICU: Intensive Care Unit
AP-HP: Assistance PubliqueHôpitaux de Paris
ICD-10: International Classification of Diseases, 10th Revision
CCAM: Classification Commune des Actes Médicaux
Insee: Institut national de la statistique et des études économiques (National Institute of Statistics and Economic Studies)
RRT: Renal Replacement Therapy
ATE: Average Treatment Effect
ATT: Average Treatment effect on the Treated
LLM: Large language models

## Acknowledgements

None.

