## Supplementary material for "Propensity-score matching with GAN-generated observations from electronic health records: simulation study and application to the evaluation of prone positioning in COVID-19 patients under mechanical ventilation"

Supplementary Text 1. Parameters of the simulation study 2

Supplementary Table 1. Specification of the protocol of a target trial and its emulation to estimate the effect of prone positioning on 28-day mortality of COVID-19 patients in respiratory distress 3

Supplementary Table 2. Characteristics of patients in the cohort extracted from the electronic health record 4

Supplementary Table 3. Characteristics of patients after multiple imputations 6

Supplementary Table 4. Thresholds values used to discretize the continuous variables 8

Supplementary Table 5. Performances of matching frameworks over 1000 simulations under the alternative hypotheses with simulated log OR = 0.5 9

Supplementary Figure 1. Comparison of the absolute Means and standard deviations (STDs) between observed and generated data. 10

Supplementary Figure 2. Comparison of the cumulative sum of every feature between observed and generated data. 11

Supplementary Figure 3. Comparison of the correlations between observed and generated data 19

Supplementary Figure 4. Visualization of the standardized mean difference of the features between the treated and the control group for each of the matching methods 20

**Supplementary Text 1.** Parameters of the simulation study

Each simulated dataset consisted of 2000 patients with 8 baseline variables: 6 binary covariates representing patient characteristics ($X_1$–$X_6$), 1 binary treatment indicator (T) and 1 binary outcome (Y) for each hypothesis.
Patient characteristics $X_1$ to $X_6$ were drawn from multivariate normal distribution using a mean of 0 for each covariate, and the following covariance matrix:

$$\begin{pmatrix} 1 & 0.6 & 0.3 & 0 & -0.2 & -0.6 \\ 0.6 & 1 & 0.6 & 0.3 & 0 & -0.2 \\ 0.3 & 0.6 & 1 & 0.6 & 0.3 & 0 \\ 0 & 0.3 & 0.6 & 1 & 0.6 & 0.3 \\ -0.2 & 0 & 0.3 & 0.6 & 1 & 0.6 \\ -0.6 & -0.2 & 0 & 0.3 & 0.6 & 1 \end{pmatrix}$$

Each covariate was then dichotomized using the median of the covariates as thresholds.

Treatment allocations and outcomes were drawn from Bernoulli distribution with the following probabilities:

$$P(T = 1 \mid X) = \frac{1}{1 + e^{-2x_1 + x_2 + 0.5\,x_3 + x_5 - 2x_6}}$$

Outcomes Y were drawn from Bernoulli laws under different hypotheses, with outcome probability calibrated so that $P(Y = 1) \approx 0.3$ under all hypotheses.

Null hypotheses H0a and H0b considered that the treatment had no effect on the outcome, with a 0.3 constant probability for H0a and a probability depending on baseline characteristics for H0b:

$$P(Y_{H0a} = 1) = 0.3 \qquad P(Y_{H0b} = 1 \mid X) = \frac{1}{1 + e^{1.4 - \frac{x_1 + x_2 - x_3 - x_4}{4}}}$$

Alternative hypotheses H1a and H1b considered that the treatment had an effect on the outcome ($log(OR) = 0.1$), with H1b implying additional associations of the outcome with baseline characteristics:

$$P(Y_{H1a} = 1 \mid T) = \frac{1}{1 + e^{0.8 - 0.1 \cdot T}} \qquad P(Y_{H1b} = 1 \mid T, X) = \frac{1}{1 + e^{1.3 - 0.1 \cdot T - \frac{x_1 + x_2 - x_3 - x_4}{4}}}$$

A sensitivity analysis was conducted with a higher effect size for H1a and H1b ($log(OR) = 0.5$):

$$P\left(Y_{H1a_2} = 1 \mid T\right) = \frac{1}{1 + e^{0.6 - 0.5 \cdot T}} \qquad P\left(Y_{H1b_2} = 1 \mid T, X\right) = \frac{1}{1 + e^{1.1 - 0.5 \cdot T - \frac{x_1 + x_2 - x_3 - x_4}{4}}}$$

**Supplementary Table 1.** Specification of the protocol of a target trial and its emulation to estimate the effect of prone positioning on 28-day mortality of COVID-19 patients in respiratory distress

| Protocol component | Target trial specification description | Target trial emulation description |
|---|---|---|
| **Eligibility criteria** | - COVID-19 positive patients<br>- age > 18 years<br>- ICU admission<br>- patients in respiratory distress early after hospitalization<br>- mechanical ventilation onset early after hospitalization<br>- patients not vaccinated | - COVID-19 positive patients<br>- age > 18 years<br>- minimum 48-hour hospital stay<br>- ICU admission with documented 28-day vital status<br>- patients in respiratory distress within 48 hours following ICU admission<br>- mechanical ventilation within 48 hours following ICU admission<br>- hospital admission between March 1, 2020 and December 31, 2020 |
| **Treatment strategies** | Comparison of two intervention groups:<br>- alternate prone position onset at ICU admission (treatment group)<br>- no prone position onset at ICU admission (control group) | Comparison of two groups:<br>- alternate prone position within 48 hours following ICU admission (treatment group)<br>- no prone position within 48 hours following ICU admission (control group) |
| **Assignment procedures** | - patients randomly assigned to an intervention group at baseline<br>- no blinding of the patients or the investigator | Patients groups defined depending on prone positioning within 48 hours following ICU admission |
| **Follow-up period** | - starts at randomization<br>- ends 28 days after randomization or if death occurs | - starts within 48 hours following ICU admission<br>- ends 28 days later or if death occurs |
| **Outcomes** | 28-day patient mortality | 28-day patient mortality |
| **Causal contrasts of interest** | - intention-to-treat effect<br>- per-protocol effect | Observational analog of intention-to-treat effect |
| **Statistical analysis** | - intention-to-treat effect: estimation using the comparison of 28-day mortality between patients of each intervention strategy<br>. adjustments for pre- and postbaseline prognostic factors associated with loss to follow-up<br><br>- per-protocol effect<br>. adjustments for pre- and postbaseline prognostic factors associated with adherence to the strategy<br>. adjustments for pre- and postbaseline prognostic factors associated with loss to follow-up | Observational analog of intention-to-treat effect: estimation based on matching on baseline covariates with a propensity score |

**Supplementary Table 2.** Characteristics of patients in the cohort extracted from the electronic health record

Continuous variables are described as median [Q1; Q3], and the discrete variables as the count of positive patients for the variable.

| | Units | Missing data | Control group n = 809 | | Prone positioning group n = 590 | |
|---|---|---|---|---|---|---|
| | | | Non-survivors n = 254 | Survivors n = 555 | Non-survivors n = 226 | Survivors n = 364 |
| Sex | - | 0% | M: 193; F: 61 | M: 414; F: 141 | M: 176; F: 50 | M: 260; F: 104 |
| Age | years | 0% | 68 [60; 74] | 61 [52; 69] | 67 [60; 72] | 62 [54; 69.25] |
| Eosinophils (blood) | $10^9$/L | 24.16% | 0.0 [0.0; 0.01] | 0 [0.0; 0.02] | 0.0 [0.0; 0.0] | 0.0 [0.0; 0.01] |
| Oxygen saturation (blood) | % | 20.01% | 95.9 [92.4; 98.0] | 95.1 [91.4; 97.8] | 92.6 [87.3; 96.3] | 94.8 [90.6; 97.3] |
| Leukocytes (blood) | $10^9$/L | 21.59% | 8.63 [5.99; 12.59] | 8.9 [6.40; 13.70] | 8 [5.95; 11.80] | 9.22 [6.66; 12.63] |
| Monocytes (blood) | $10^9$/L | 24.16% | 0.45 [0.24; 0.72] | 0.41 [0.28; 0.68] | 0.365 [0.22; 0.60] | 0.36 [0.23; 0.56] |
| Bilirubin total (serum) | µmol/L | 20.37% | 10.0 [7.0; 15.0] | 9.2 [7.0; 14.0] | 9.0 [7.0; 14.0] | 9.0 [7.0; 13.0] |
| Prothrombin Ratio | % | 15.08% | 75.5 [65.0; 88.2] | 82.0 [70.0; 93.0] | 80.0 [69.0; 91.0] | 83.0 [73.0; 93.0] |
| Lymphocytes (blood) | $10^9$/L | 24.16% | 0.81 [0.50; 1.29] | 0.88 [0.61; 1.24] | 0.75 [0.50; 1.03] | 0.87 [0.63; 1.22] |
| Platelets (blood) | $10^9$/L | 21.73% | 191 [148; 278] | 228 [170; 304] | 196 [141; 251] | 228 [175; 292] |
| Sodium (serum) | mmol/L | 12.44% | 136 [133; 139] | 136 [134; 139] | 135 [133; 138] | 136 [133; 139] |
| Neutrophils (blood) | $10^9$/L | 24.09% | 7.11 [4.8; 10.49] | 7.43 [5.14; 11.93] | 6.89 [4.55; 10.53] | 7.76 [5.39; 10.39] |
| Hemoglobin (blood) | g/dL | 21.66% | 12.4 [10.3; 13.8] | 12.8 [11.3; 14.3] | 12.9 [11.7; 14.1] | 12.8 [11.3; 14] |
| Oxygen pressure adjusted to the patient temperature | mmHg | 24.8% | 82.1 [65.9; 112.0] | 79.0 [63.4; 105.0] | 69.9 [55.4; 90.7] | 75.0 [61.2; 97.0] |
| Carbon dioxide pressure corrected by temperature (arterial blood) | mmHg | 20.59% | 37.0 [32.4; 46.4] | 38.1 [33.5; 44.225] | 37.3 [33.3; 42.5] | 37.2 [33; 43.05] |
| pH adjusted to the patient temperature (arterial blood) | - | 17.58% | 7.40 [7.31; 7.44] | 7.43 [7.37 7.47] | 7.42 [7.35; 7.46] | 7.43 [7.37; 7.46] |
| Bicarbonate (arterial blood) | mmol/L | 24.3% | 23.3 [20.2; 26.1] | 24.6 [22.0; 27.4] | 23.9 [21.6; 26.1] | 23.6 [22.0; 26.2] |
| Potassium (serum) | mmol/L | 25.88% | 4.1 [3.8; 4.5] | 4.0 [3.6; 4.4] | 4.0 [3.7; 4.4] | 3.9 [3.5; 4.3] |
| Lactate (blood) | mmol/L | 27.31% | 1.6 [1.1; 2.4] | 1.4 [1.1; 1.9] | 1.5 [1.1; 2.1] | 1.4 [1.0; 1.9] |
| Basophils (blood) | $10^9$/L | 24.16% | 0.01 [0.0; 0.02] | 0.01 [0.0; 0.02] | 0.01 [0.0; 0.02] | 0.01 [0; 0.02] |
| Urea (serum) | mmol/L | 17.94% | 8.5 [5.8; 14.7] | 6.4 [4.7; 9.1] | 7.7 [6.0; 13.0] | 6.4 [4.6; 9.8] |
| Activated partial thromboplastin time patient/control | - | 18.08% | 1.16 [1.05; 1.37] | 1.16 [1.03; 1.32] | 1.17 [1.03; 1.31] | 1.164 [1.04; 1.30] |
| Fibrinogen activity (Clauss method) | g/L | 19.44% | 5.9 [4.9; 7.6] | 6.5 [5.3; 7.6] | 6.3 [5.3; 7.6] | 7.1 [6.0; 8.1] |
| Respiratory diseases: pneumonia | | 0% | 117 (46%) | 271 (49%) | 146 (65%) | 222 (61%) |
| Respiratory distress | | 0% | 186 (73%) | 384 (69%) | 198 (88%) | 308 (85%) |
| Septic shock | | 0% | 79 (31%) | 101 (18%) | 97 (43%) | 84 (23%) |
| Isolation | | 0% | 105 (41%) | 258 (46%) | 128 (57%) | 237 (65%) |
| Hypertension | | 0% | 86 (34%) | 155 (28%) | 91 (40%) | 143 (39%) |
| Renal diseases | | 0% | 129 (51%) | 116 (21%) | 122 (54%) | 96 (26%) |

| | | | | | |
|---|---|---|---|---|---|
| Systemic inflammatory response | 0% | 50 (20%) | 103 (19%) | 59 (26%) | 64 (18%) |
| Hepatic insufficiency | 0% | 15 (6%) | 10 (2%) | 9 (4%) | 6 (2%) |
| Heart failure | 0% | 30 (12%) | 35 (6%) | 10 (4%) | 15 (4%) |
| Obesity / overweight | 0% | 55 (22%) | 133 (24%) | 68 (30%) | 115 (32%) |
| Diabetes | 0% | 69 (27%) | 100 (18%) | 66 (29%) | 96 (26%) |
| Cancer | 0% | 25 (10%) | 34 (6%) | 16 (7%) | 15 (4%) |
| Disorientation / Symptoms related to cognitive functions and consciousness | 0% | 6 (2%) | 25 (5%) | 8 (4%) | 14 (4%) |
| Asthma | 0% | 6 (2%) | 28 (5%) | 7 (3%) | 11 (3%) |
| Dobutamine / dopamine injection | 0% | 193 (76%) | 357 (64%) | 182 (81%) | 242 (66%) |
| Intravenous infusion filler | 0% | 83 (33%) | 123 (22%) | 83 (37%) | 105 (29%) |
| Intra-arterial pressure monitoring | 0% | 198 (78%) | 424 (76%) | 175 (77%) | 277 (76%) |
| Central venous catheter | 0% | 157 (62%) | 338 (61%) | 146 (65%) | 254 (70%) |
| Continuous sedation / curarization | 0% | 65 (26%) | 171 (31%) | 80 (35%) | 153 (42%) |
| Enteral feeding by tube | 0% | 116 (46%) | 272 (49%) | 138 (61%) | 200 (55%) |
| Tracheal intubation | 0% | 119 (47%) | 260 (47%) | 113 (50%) | 196 (54%) |
| Extracorporeal membrane oxygenation | 0% | 21 (8%) | 53 (10%) | 9 (4%) | 6 (2%) |
| Renal replacement therapy | 0% | 1 (0.4%) | 2 (0.4%) | 0 (0%) | 0 (0%) |
| Corticosteroids administration | 0% | 5 (2%) | 10 (2%) | 2 (1%) | 6 (6%) |

**Supplementary Table 3.** Characteristics of patients after multiple imputations

Continuous variables are described as median [Q1; Q3], and the discrete variables as the count of positive patients for the variable.

| | Units | Control group n = 809 | | Prone positioning group n = 590 | |
|---|---|---|---|---|---|
| | | **Non-survivors** n = 254 | **Survivors** n = 555 | **Non-survivors** n = 226 | **Survivors** n = 364 |
| Sex | - | M: 193; F: 61 | M: 414; F: 141 | M: 176; F: 50 | M: 260; F: 104 |
| Age | years | 68 [60; 74] | 61 [52; 69] | 67 [60; 72] | 62 [54; 69] |
| Eosinophils (blood) | $10^9$/L | 0.0 [0.0; 0.01] | 0.0 [0.0; 0.02] | 0.0 [0.0; 0.0] | 0.0 [0.0; 0.01] |
| Oxygen saturation (blood) | % | 95.6 [92.0; 98.0] | 95.1 [91.2; 97.8] | 92.5 [87.3; 96.3] | 94.9 [90.8; 97.5] |
| Leukocytes (blood) | $10^9$/L | 9.04 [6.30; 13.28] | 8.76 [6.23; 13.60] | 8.28 [5.98; 11.99] | 9.06 [6.54; 12.70] |
| Monocytes (blood) | $10^9$/L | 0.49 [0.28; 0.79] | 0.43 [0.28; 0.7] | 0.38 [0.23; 0.61] | 0.38 [0.24; 0.59] |
| Bilirubin total (serum) | µmol/L | 10 [7; 15] | 9.2 [7; 14] | 9 [6.5; 13] | 9 [7; 13] |
| Prothrombin Ratio | % | 75 [64; 88] | 82 [71; 93] | 80.5 [68; 91] | 84 [74; 94] |
| Lymphocytes (blood) | $10^9$/L | 0.83 [0.51; 1.36] | 0.88 [0.6; 1.26] | 0.76 [0.52; 1.10] | 0.87 [0.6; 1.22] |
| Platelets (blood) | $10^9$/L | 198.0 [148.0; 286.7] | 226.0 [169.0; 297.5] | 202.5 [150.5; 261.7] | 228.0 [172.0; 293.2] |
| Sodium (serum) | mmol/L | 136 [133; 140] | 136 [134; 138] | 135 [133; 138] | 136 [133; 139] |
| Neutrophils (blood) | $10^9$/L | 7.42 [5.07; 10.83] | 7.18 [4.89; 10.96] | 7.10 [4.58; 10.81] | 7.70 [5.32; 10.72] |
| Hemoglobin (blood) | g/dL | 12.3 [10.1; 13.8] | 12.9 [11.3; 14.3] | 12.8 [11.6; 14.0] | 12.9 [11.3; 14.1] |
| Oxygen pressure adjusted to the patient temperature | mmHg | 82.1 [66.0; 111.5] | 79.5 [63.7; 104.5] | 70.85 [57.6; 91.8] | 76.0 [62.2; 100.6] |
| Carbon dioxide pressure corrected by temperature (arterial blood) | mmHg | 36.9 [32.8; 45.5] | 38.0 [33.3; 43.2] | 36.3 [33.0; 42.3] | 36.4 [33.0; 42.9] |
| pH adjusted to the patient temperature (arterial blood) | - | 7.39 [7.31; 7.45] | 7.43 [7.37; 7.47] | 7.42 [7.35; 7.46] | 7.43 [7.37; 7.46] |
| Bicarbonate (arterial blood) | mmol/L | 23.2 [20.0; 26.1] | 24.5 [22.0; 27.3] | 23.6 [21.4; 26.0] | 23.8 [22.0; 26.3] |
| Potassium (serum) | mmol/L | 4.1 [3.7; 4.5] | 4.0 [3.6; 4.4] | 4.0 [3.7; 4.4] | 3.9 [3.5; 4.3] |
| Lactate (blood) | mmol/L | 1.6 [1.1; 2.5] | 1.4 [1.1; 1.9] | 1.5 [1.1; 2.1] | 1.4 [1.1; 2] |
| Basophils (blood) | $10^9$/L | 0.01 [0; 0.02] | 0.01 [0; 0.02] | 0.01 [0; 0.02] | 0.01 [0.01; 0.02] |
| Urea (serum) | mmol/L | 8.5 [5.8; 14.6] | 6.4 [4.8; 9.1] | 7.6 [5.8; 12.4] | 6.6 [4.7; 9.8] |
| Activated partial thromboplastin time patient/control | - | 1.17 [1.05; 1.37] | 1.17 [1.04; 1.33] | 1.16 [1.03; 1.3] | 1.17 [1.05; 1.31] |
| Fibrinogen activity (Clauss method) | g/L | 5.89 [4.87; 7.43] | 6.5 [5.25; 7.45] | 6.49 [5.30; 7.77] | 7.06 [6.00; 7.99] |
| Respiratory diseases: pneumonia | | 117 (46%) | 271 (49%) | 146 (65%) | 222 (61%) |
| Respiratory distress | | 186 (73%) | 384 (69%) | 198 (88%) | 308 (85%) |
| Septic shock | | 79 (31%) | 101 (18%) | 97 (43%) | 84 (23%) |
| Isolation | | 105 (41%) | 258 (46%) | 128 (57%) | 237 (65%) |
| Hypertension | | 86 (34%) | 155 (28%) | 91 (40%) | 143 (39%) |

| | | | | |
|---|---|---|---|---|
| Renal diseases | 129 (51%) | 116 (21%) | 122 (54%) | 96 (26%) |
| Systemic inflammatory response | 50 (20%) | 103 (19%) | 59 (26%) | 64 (18%) |
| Hepatic insufficiency | 15 (6%) | 10 (2%) | 9 (4%) | 6 (2%) |
| Heart failure | 30 (12%) | 35 (6%) | 10 (4%) | 15 (4%) |
| Obesity / overweight | 55 (22%) | 133 (24%) | 68 (30%) | 115 (32%) |
| Diabetes | 69 (27%) | 100 (18%) | 66 (29%) | 96 (26%) |
| Cancer | 25 (10%) | 34 (6%) | 16 (7%) | 15 (4%) |
| Disorientation / Symptoms related to cognitive functions and consciousness | 6 (2%) | 25 (5%) | 8 (4%) | 14 (4%) |
| Asthma | 6 (2%) | 28 (5%) | 7 (3%) | 11 (3%) |
| Dobutamine / dopamine injection | 193 (76%) | 357 (64%) | 182 (81%) | 242 (66%) |
| Intravenous infusion filler | 83 (33%) | 123 (22%) | 83 (37%) | 105 (29%) |
| Intra-arterial pressure monitoring | 198 (78%) | 424 (76%) | 175 (77%) | 277 (76%) |
| Central venous catheter | 157 (62%) | 338 (61%) | 146 (65%) | 254 (70%) |
| Continuous sedation / curarization | 65 (26%) | 171 (31%) | 80 (35%) | 153 (42%) |
| Enteral feeding by tube | 116 (46%) | 272 (49%) | 138 (61%) | 200 (55%) |
| Tracheal intubation | 119 (47%) | 260 (47%) | 113 (50%) | 196 (54%) |
| Extracorporeal membrane oxygenation | 21 (8%) | 53 (10%) | 9 (4%) | 6 (2%) |
| Renal replacement therapy | 1 (0.4%) | 2 (0.4%) | 0 (0%) | 0 (0%) |
| Corticosteroids administration | 5 (2%) | 10 (2%) | 2 (1%) | 6 (6%) |

**Supplementary Table 4.** Thresholds values used to discretize the continuous variables
F: female patients, M: male patients.

| Continuous variables | Units | Threshold values |
|---|---|---|
| Age | years | 60; 70 |
| Eosinophils blood | $10^9$/L | 0.5 |
| Oxygen saturation blood | % | 95 |
| Leukocytes blood | $10^9$/L | 4;<br>10 |
| Monocytes blood | $10^9$/L | 0.2; 1 |
| Bilirubin total serum plasma | µmol/L | 20 |
| Prothrombin Ratio | % | 70 |
| Lymphocytes blood | $10^9$/L | 1; 4 |
| Platelets blood | $10^9$/L | 150; 450 |
| Sodium serum plasma | mmol/L | 135; 145 |
| Neutrophils blood | $10^9$/L | 1.5; 70 |
| Hemoglobin blood automated counting | g/dL | 12 (F); 13 (M) |
| Oxygen pressure adjusted to the patient temperature | mmHg | 90 |
| Carbon dioxide pressure corrected by temperature blood | mmHg | 35; 45 |
| Ph adjusted to the patient temperature | - | 7.38; 7.42 |
| Bicarbonate blood arterial | mmol/L | 22; 28 |
| Potassium serum plasma | mmol/L | 3.5; 4.5 |
| Lactate blood specific electrodes | mmol/L | 20 |
| Basophils blood | $10^9$/L | 0.1 |
| Urea serum plasma | mmol/L | 7 (F); 7.5 (M) |
| Activated partial thromboplastin time patient/control | - | 0.8; 1.2 |
| Fibrinogen activity (Clauss method) | g/L | 4; 22 |

**Supplementary Table 5.** Performances of matching frameworks over 1000 simulations under the alternative hypotheses with simulated log OR = 0.5

H1a and H1b: absence and presence of confounding, respectively. PSM: propensity-score matching of simulated patients (no GAN-generated patients). PHM: partial hybrid (PSM with additional “hybrid pairs” of simulated and GAN-generated synthetic patients). FHM: full hybrid (matching of simulated patients with synthetic ones). Probability of treatment allocation set to either 50% (balanced) or 20% (unbalanced). Bias: average difference between simulated and estimated log OR. Empirical standard error: standard deviation of the log OR point estimates. Power: proportion of simulations with 95% CIs not containing the simulated log OR.

| | | | **H1a** | | | | | | | | **H1b** | | | | | | | |
|---|---|---|---|---|---|---|---|---|---|---|---|---|---|---|---|---|---|---|
| **Framework** | **Treatment allocation** | **Weighting of hybrid pairs** | **Total pairs** | **Synth-treated/ sim-control pairs** | **Sim-treated/ synth-control pairs** | **Bias** | **Average standard error** | **Empirical standard error** | **Power** | **95% CI coverage** | **Total pairs** | **Synth-treated/ sim-control pairs** | **Sim-treated/ synth-control pairs** | **Bias** | **Average standard error** | **Empirical standard error** | **Power** | **95% CI coverage** |
| **PSM** | **Balanced** | **-** | 711.3 | - | - | 0.006 | 0.117 | 0.119 | 0.987 | 0.946 | 711.3 | - | - | 0.026 | 0.132 | 0.132 | 0.934 | 0.938 |
| | **Unbalanced** | **-** | 398.4 | - | - | -0.007 | 0.157 | 0.162 | 0.888 | 0.946 | 398.4 | - | - | 0.000 | 0.175 | 0.175 | 0.794 | 0.957 |
| **PHM** | **Balanced** | **0.5** | 1288.7 | 288.7 | 288.8 | 0.012 | 0.098 | 0.117 | 0.993 | 0.892 | 1288.6 | 288.6 | 288.8 | 0.042 | 0.110 | 0.132 | 0.968 | 0.873 |
| | | **0.25** | 1288.7 | 288.7 | 288.8 | 0.010 | 0.106 | 0.113 | 0.996 | 0.931 | 1288.6 | 288.6 | 288.8 | 0.036 | 0.120 | 0.127 | 0.958 | 0.919 |
| | **Unbalanced** | **0.5** | 1601.3 | 1202.2 | 0.7 | 0.031 | 0.098 | 0.195 | 0.923 | 0.671 | 1601.4 | 1202.2 | 0.7 | 0.109 | 0.109 | 0.220 | 0.772 | 0.594 |
| | | **0.25** | 1601.3 | 1202.2 | 0.7 | 0.021 | 0.118 | 0.169 | 0.932 | 0.815 | 1601.4 | 1202.2 | 0.7 | 0.080 | 0.131 | 0.192 | 0.786 | 0.786 |
| **FHM** | **Balanced** | **0.5** | 2000.0 | 999.9 | 1000.0 | 0.016 | 0.098 | 0.161 | 0.970 | 0.757 | 1999.9 | 999.8 | 1000.0 | 0.059 | 0.108 | 0.174 | 0.884 | 0.748 |
| | | **0.25** | 2000.0 | 999.9 | 1000.0 | 0.016 | 0.138 | 0.161 | 0.910 | 0.908 | 1999.9 | 999.8 | 1000.0 | 0.059 | 0.153 | 0.174 | 0.767 | 0.895 |
| | **Unbalanced** | **0.5** | 1999.7 | 1600.6 | 399.1 | 0.025 | 0.098 | 0.224 | 0.897 | 0.607 | 1999.8 | 1600.6 | 399.2 | 0.105 | 0.109 | 0.253 | 0.756 | 0.538 |
| | | **0.25** | 1999.7 | 1600.6 | 399.1 | 0.025 | 0.139 | 0.224 | 0.819 | 0.777 | 1999.8 | 1600.6 | 399.2 | 0.105 | 0.154 | 0.253 | 0.611 | 0.718 |

**Supplementary Figure 1.** Comparison of the absolute Means and standard deviations (STDs) between observed and generated data. The value in the bottom left corner corresponds to renal replacement therapy, for which the GAN model produced only one value due to a small number of occurrences in the extracted cohort.

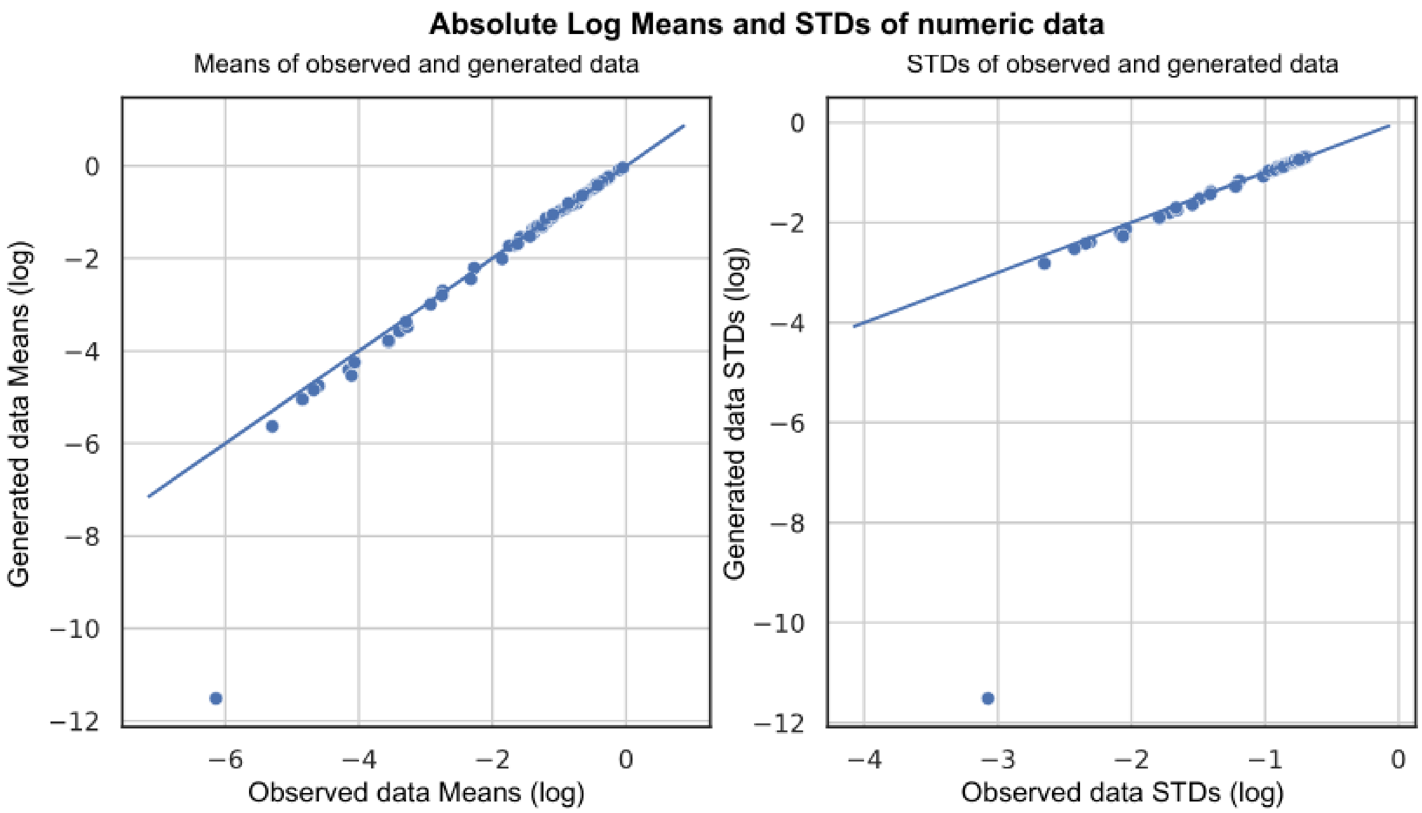

**Supplementary Figure 2.** Comparison of the cumulative sum of every feature between observed and generated data.
The GAN model produced only one value for the renal replacement therapy variable due to a small number of occurrences of this feature in the extracted cohort.

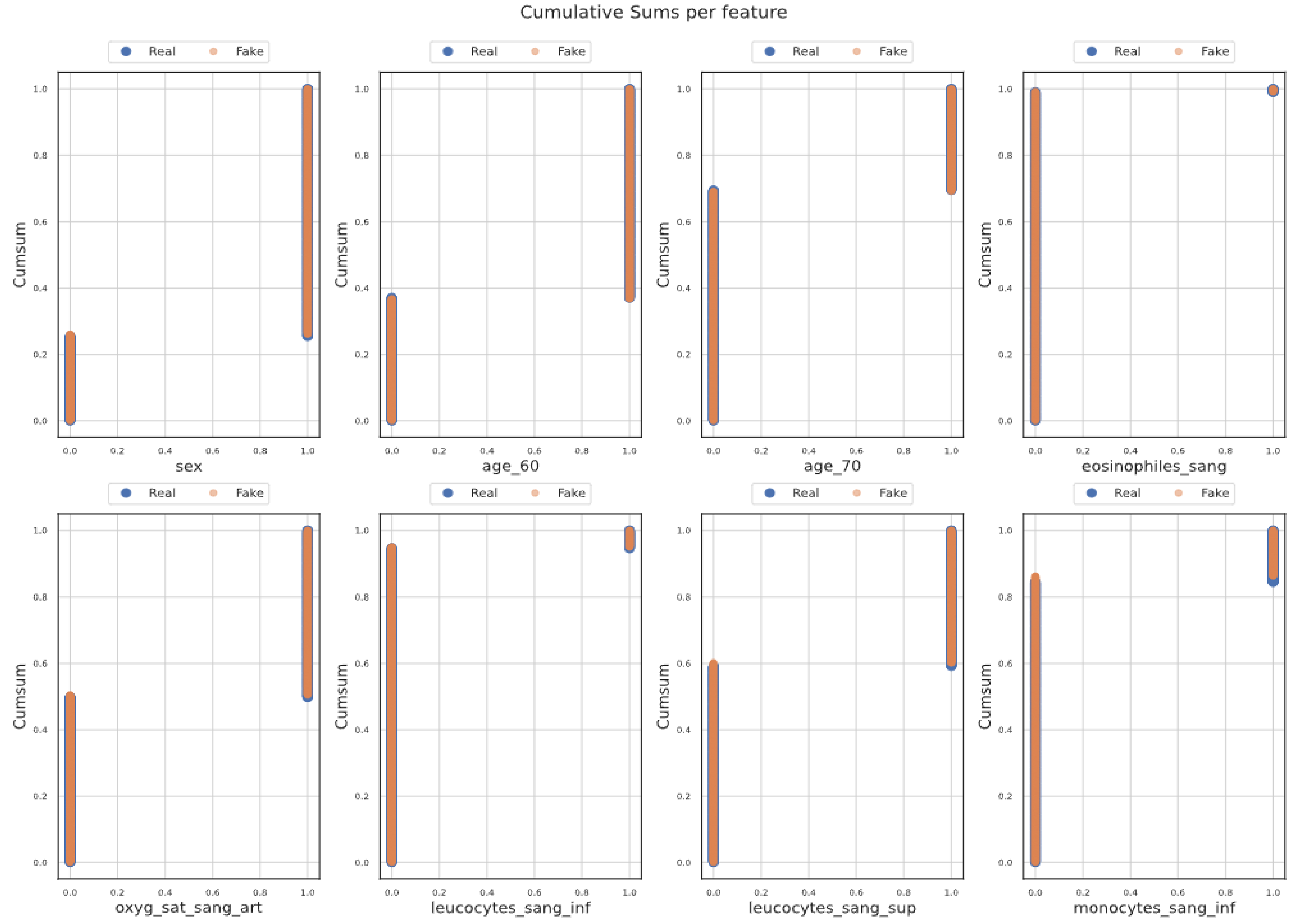

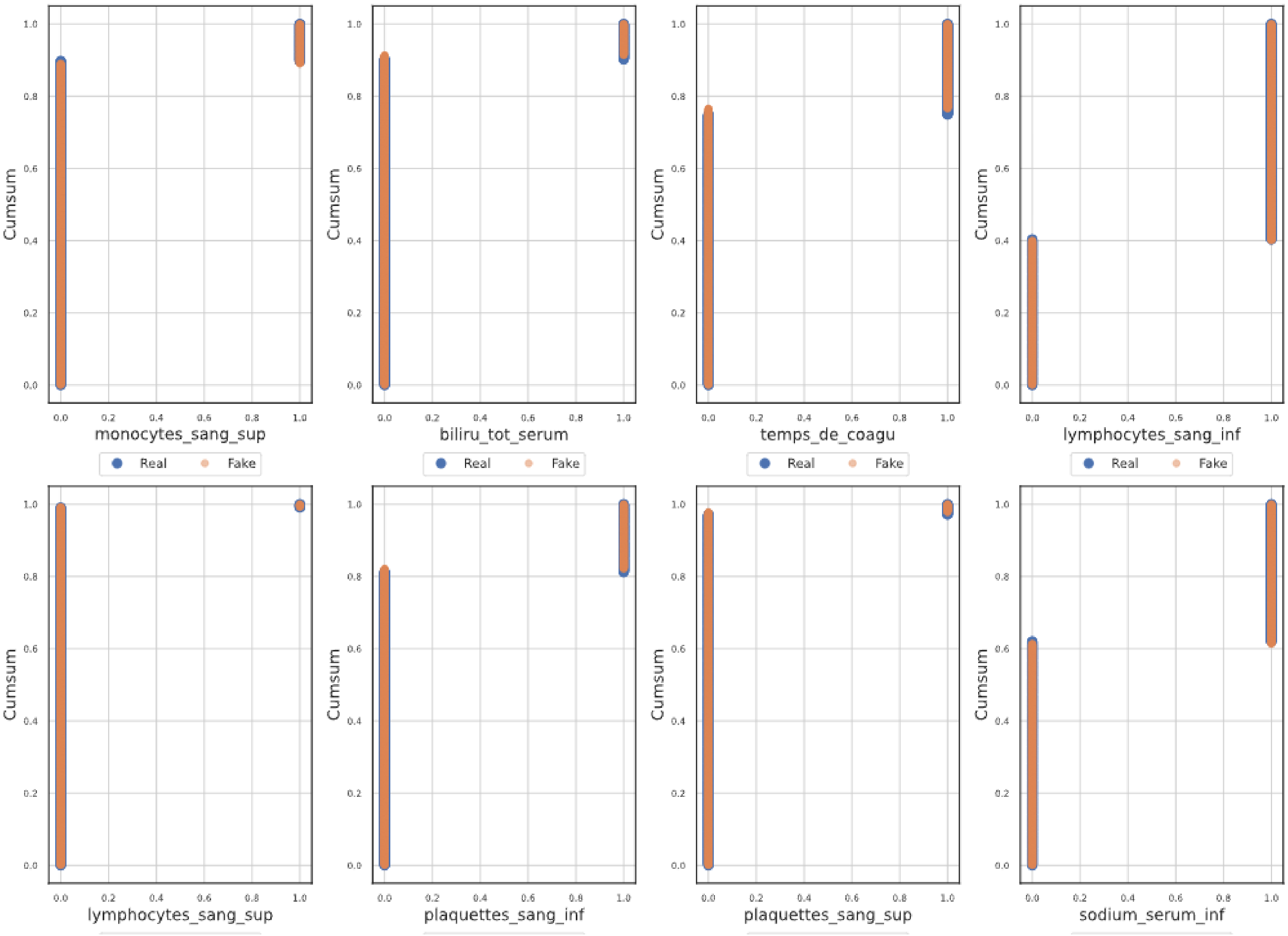
Cumsum
monocytes_sang_sup
biliru_tot_serum
temps_de_coagu
lymphocytes_sang_inf
Real
Fake
lymphocytes_sang_sup
plaquettes_sang_inf
plaquettes_sang_sup
sodium_serum_inf
0.0
0.2
0.4
0.6
0.8
1.0

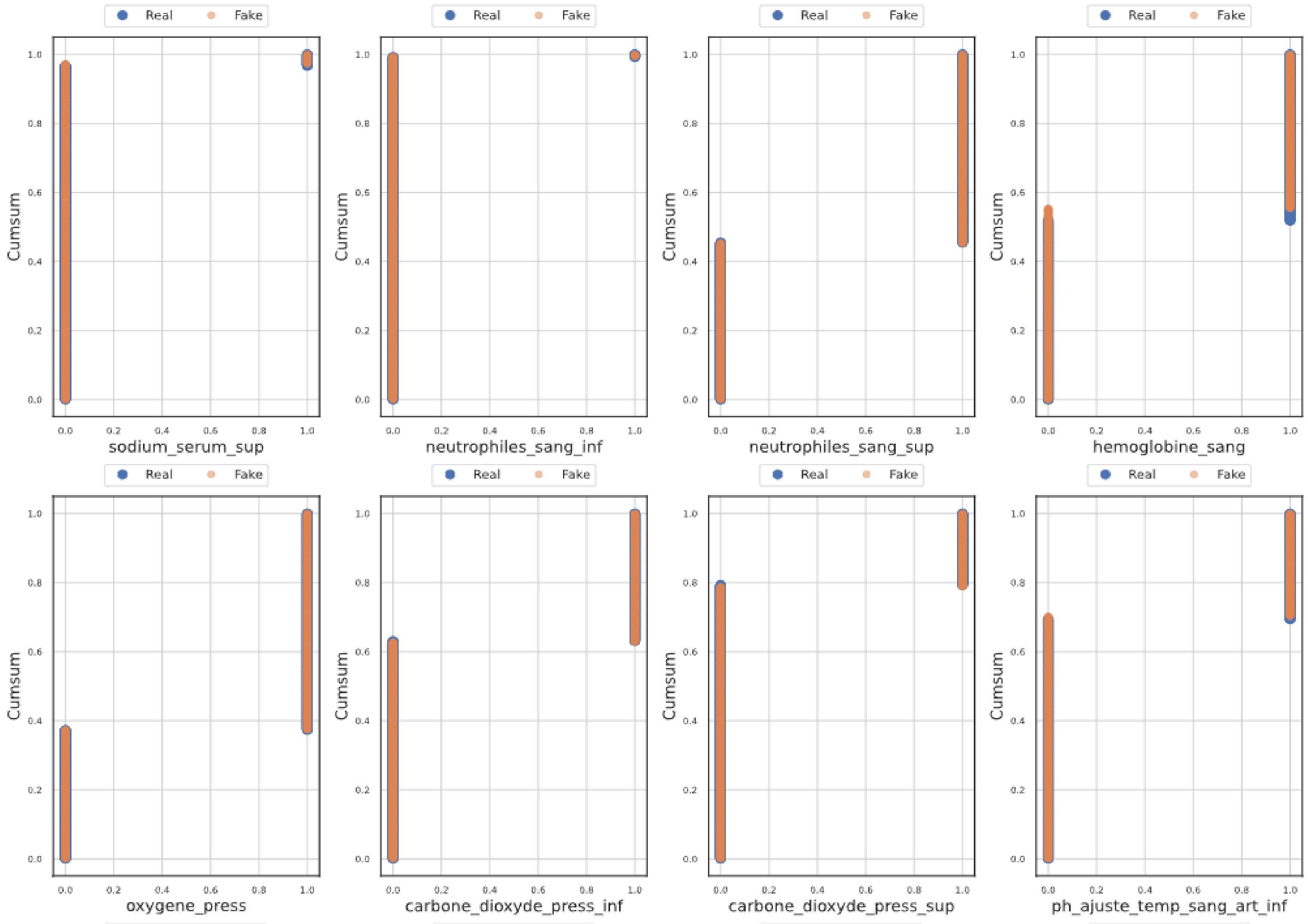
Real
Fake
Cumsum
sodium_serum_sup
neutrophiles_sang_inf
neutrophiles_sang_sup
hemoglobine_sang
oxygene_press
carbone_dioxyde_press_inf
carbone_dioxyde_press_sup
ph_ajuste_temp_sang_art_inf

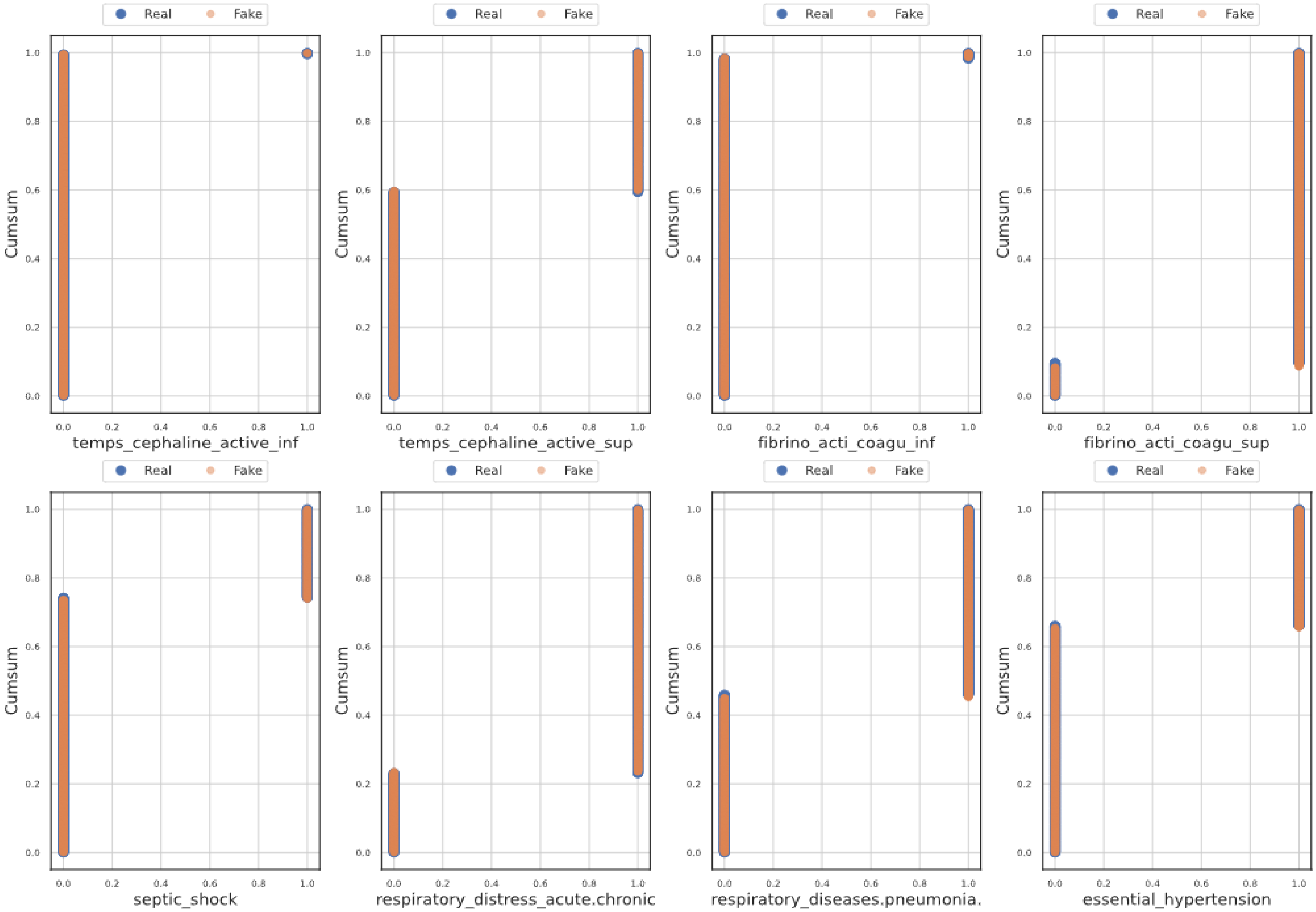
Real
Fake
Cumsum
temps_cephaline_active_inf
temps_cephaline_active_sup
fibrino_acti_coagu_inf
fibrino_acti_coagu_sup
septic_shock
respiratory_distress_acute.chronic
respiratory_diseases.pneumonia.
essential_hypertension

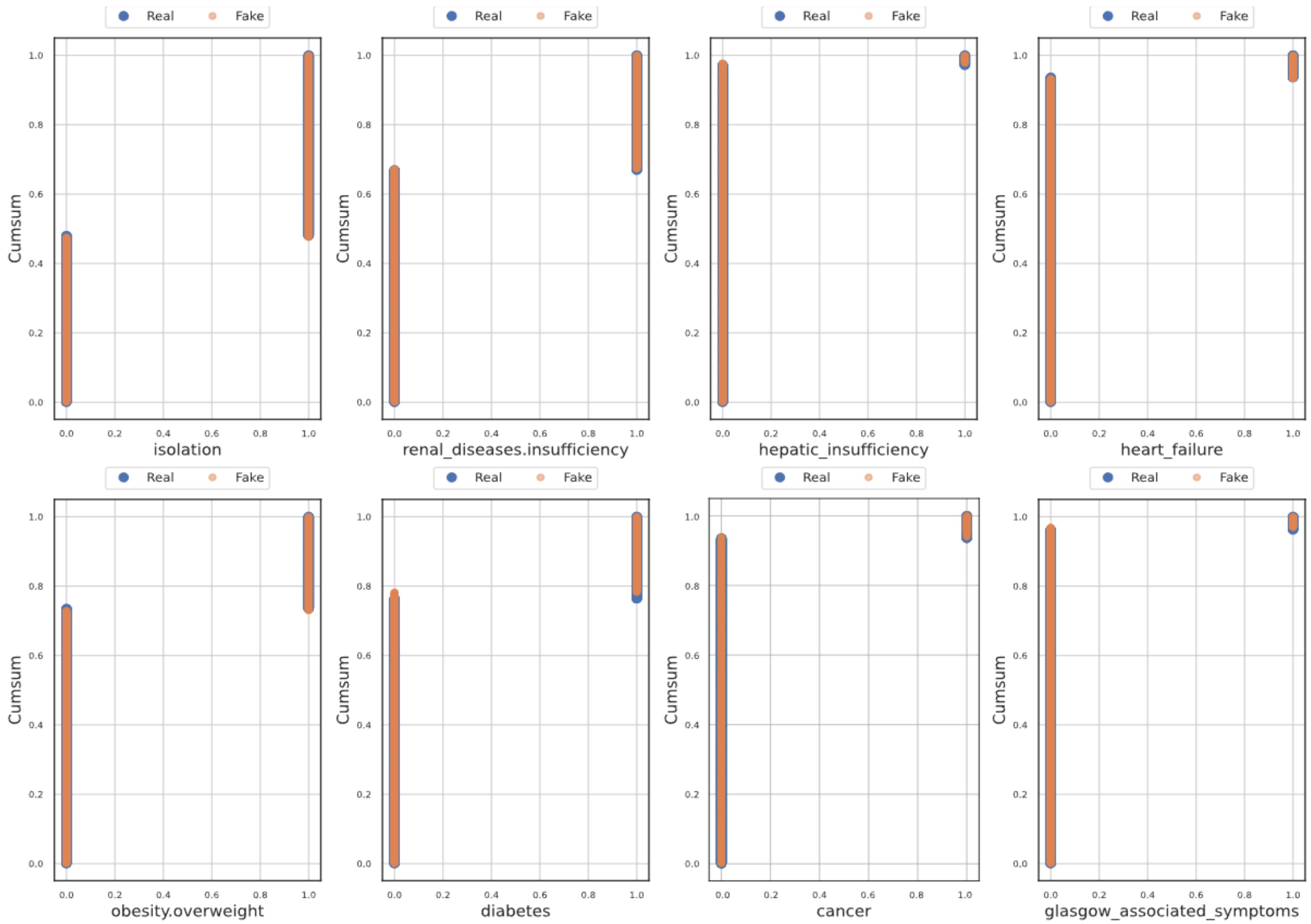
Real
Fake
Cumsum
isolation
renal_diseases.insufficiency
hepatic_insufficiency
heart_failure
obesity.overweight
diabetes
cancer
glasgow_associated_symptoms

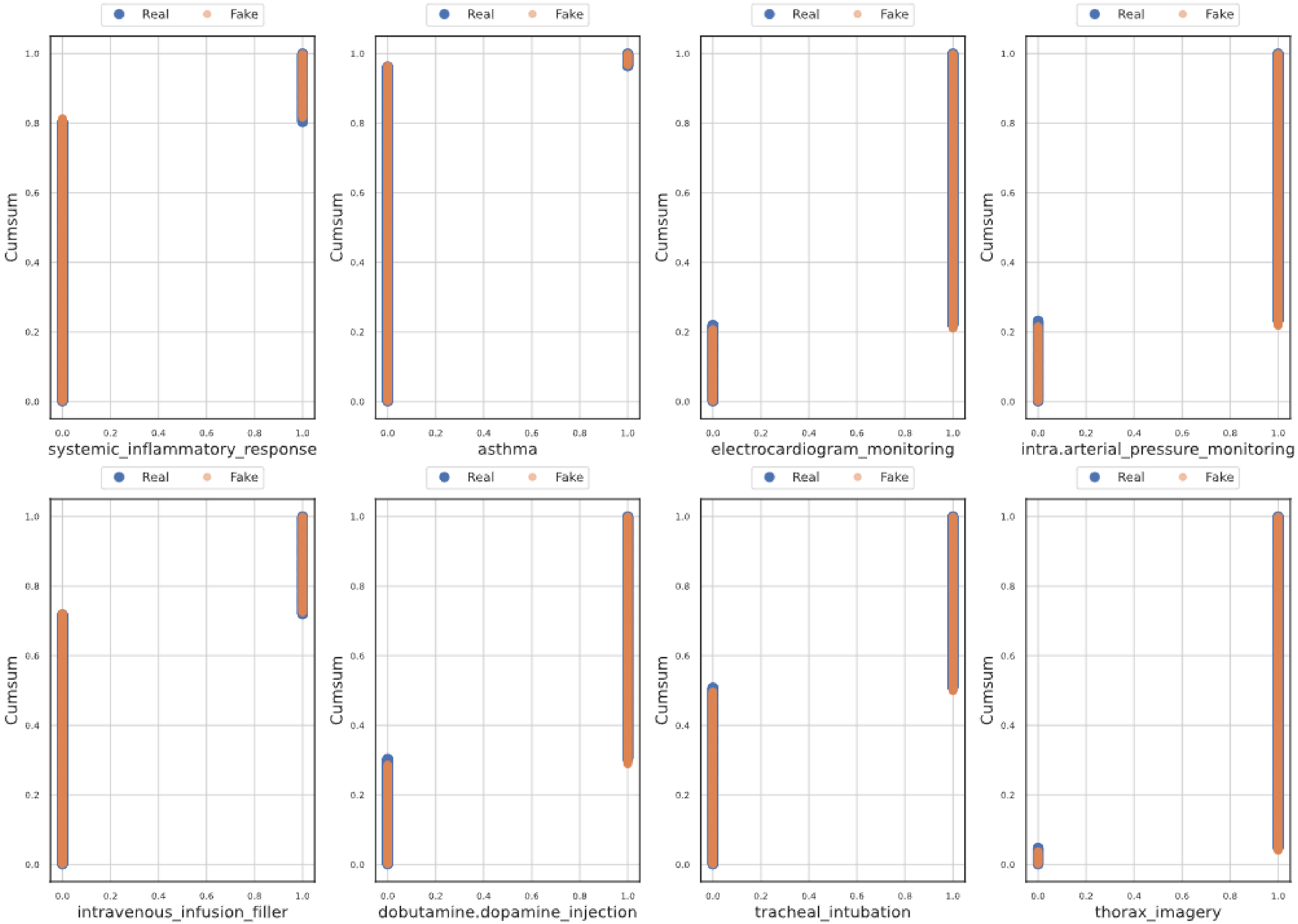
Real
Fake
Cumsum
systemic_inflammatory_response
asthma
electrocardiogram_monitoring
intra.arterial_pressure_monitoring
intravenous_infusion_filler
dobutamine.dopamine_injection
tracheal_intubation
thorax_imagery

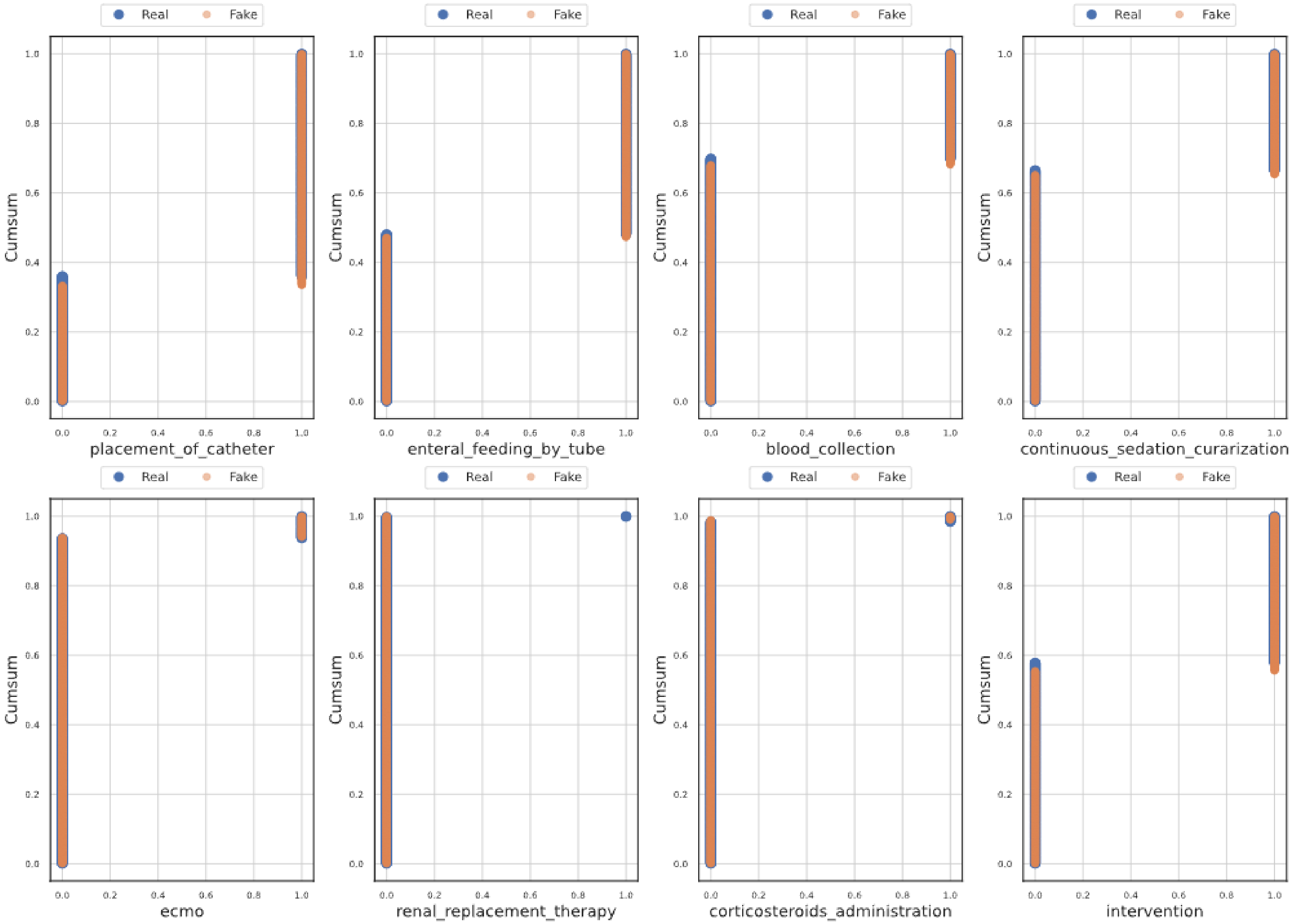
Real
Fake
Cumsum
placement_of_catheter
enteral_feeding_by_tube
blood_collection
continuous_sedation_curarization
ecmo
renal_replacement_therapy
corticosteroids_administration
intervention

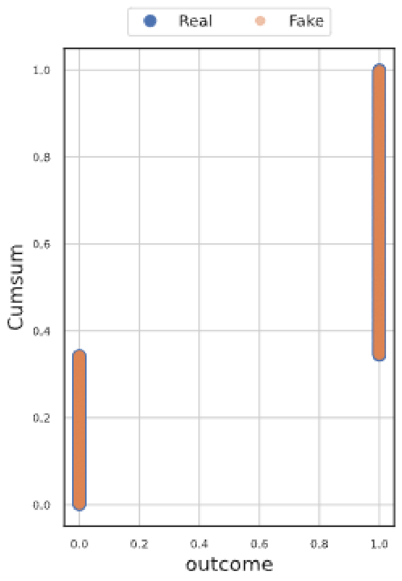

Real
Fake
1.0
0.8
0.6
0.4
0.2
0.0
Cumsum
0.0
0.2
0.4
0.6
0.8
1.0
outcome

**Supplementary Figure 3.** Comparison of the correlations between observed and generated data

Observed: correlations between variables in observed patients, Generated: correlations between variables in generated patients. Each row/column corresponds to a variable in the dataset. The gray line in the second matrix corresponds to renal replacement therapy (RRT) for which the GAN model produced only one value due to a small number of patients with RRT in observed data.

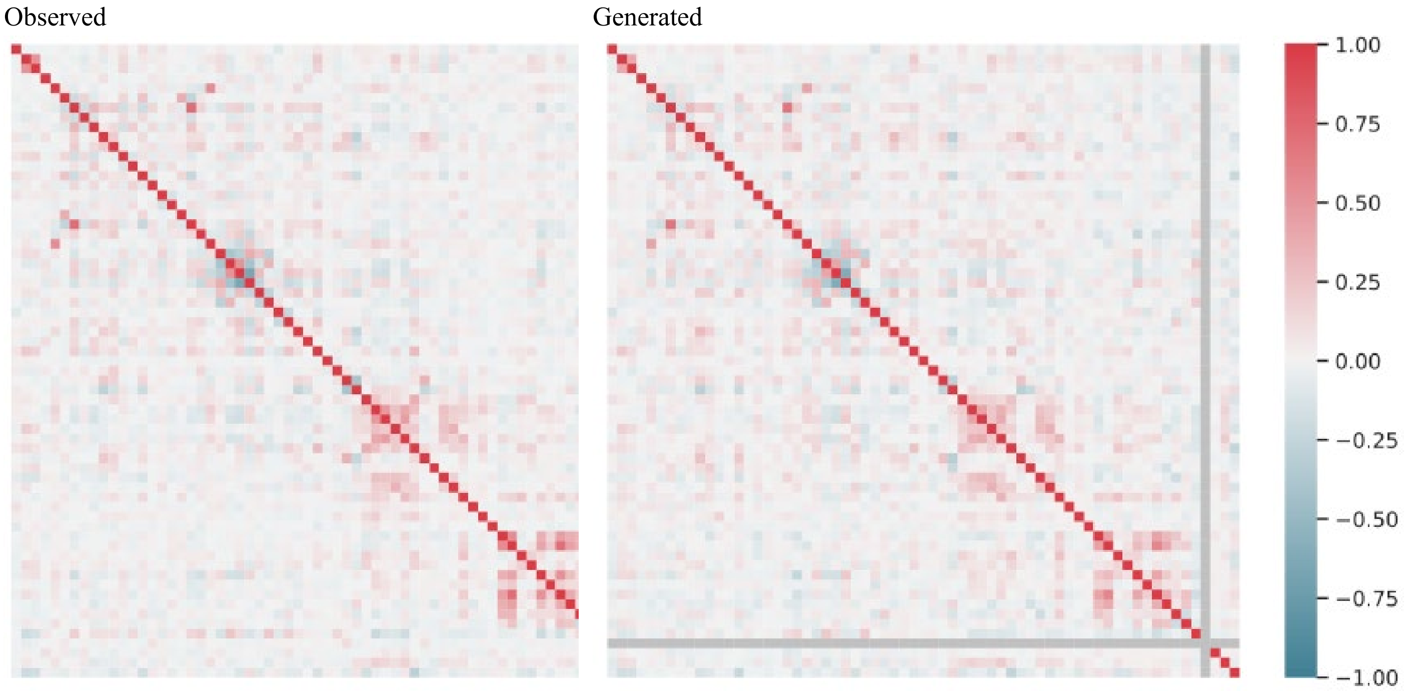

**Supplementary Figure 4.** Visualization of the standardized mean difference of the features between the treated and the control group for each of the matching methods

Unadjusted: dataset before any matching, PSM: dataset after propensity score matching, FH: dataset after full hybrid matching, PH: dataset after partial hybrid matching, SMD: standardized mean difference.

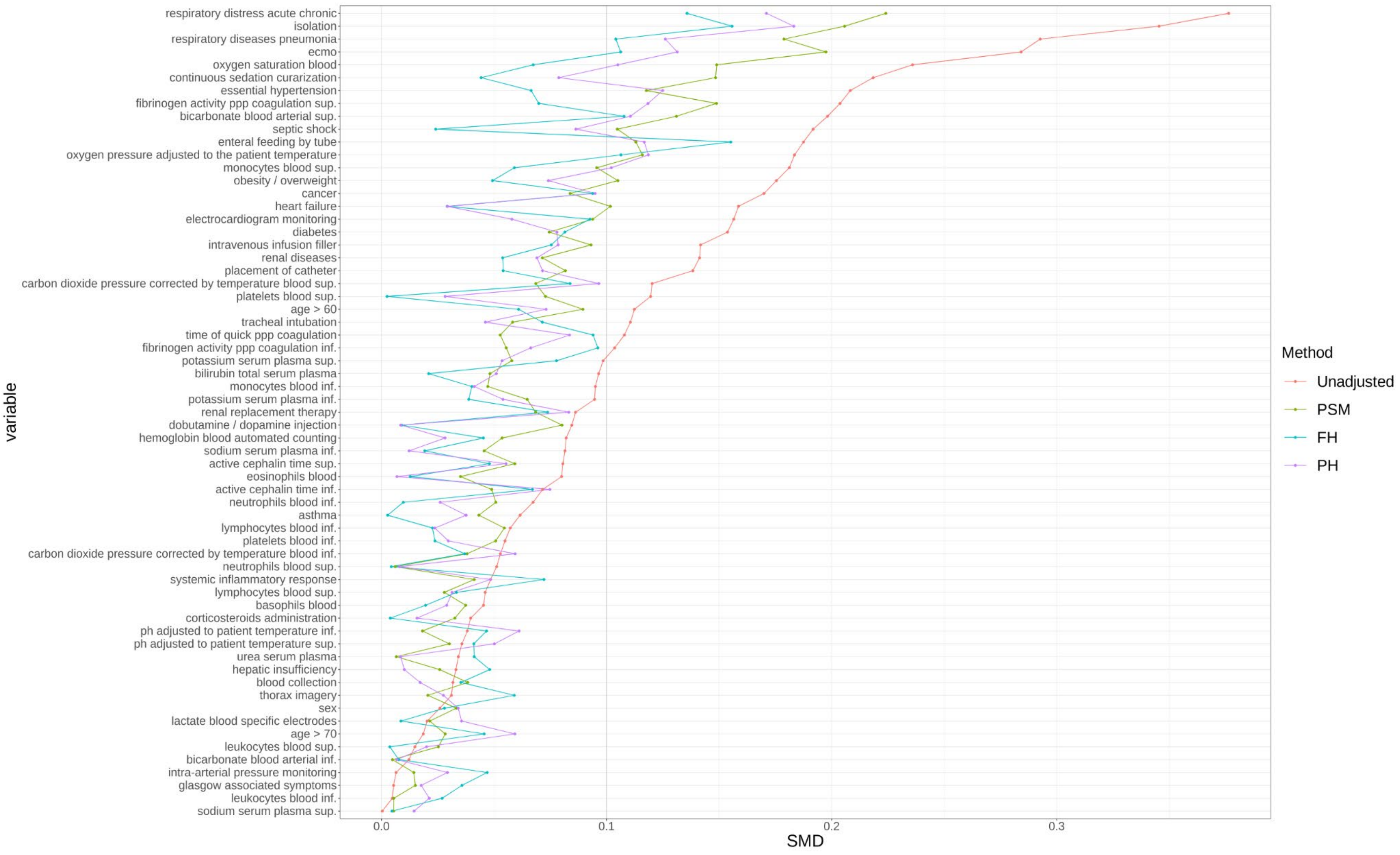
